# Molecular Epidemiology and Variant Spectrum of Rare Genetic Diseases in the Punjabi Population: A Genomic Perspective from South Asia

**DOI:** 10.1101/2025.11.05.25339593

**Authors:** Iqra Tabassum, Muhammad Shafique, Muhammad Shoaib Akhtar

## Abstract

**Background:** Rare genetic diseases (RGDs) affect individuals, families, and healthcare systems worldwide. Population-scale genomic data remain largely restricted to Western cohorts with estimated 10,000 RGDs. South Asian populations remain underrepresented in molecular, clinical, and genomic databases. This study presents the first molecular epidemiological analysis of RGDs in the Punjabi population of Pakistan.

**Methods:** Data were collected from the provincial RGD registry at the Punjab Thalassemia and Genetic Disorders Prevention and Research Institute (PTGDPRI), Lahore. Families diagnosed using next-generation sequencing (NGS) between 2021 and 2023 were enrolled. Structured questionnaires captured clinical, demographic, and socioeconomic information, and statistical and genetic analyses were performed to assess the inheritance patterns, allele frequencies, and disease distribution.

**Results:** The registry included 167 families with 72 distinct RGDs, with a mean burden of 0.81±0.24 affected children per family. Niemann–Pick disease (NP), progressive familial intrahepatic cholestasis (PFIC), and mucopolysaccharidosis (MPS) were the most common diseases. Consanguinity was observed in 89% of families, 77% of which involved first-cousin marriages, and was significantly associated with RGD incidence. Most families belonged to low-income groups despite high literacy rates, underscoring inequity in healthcare. The primary and secondary variants included 131 variants including CNV and SNVs annotated pathogenic, likely pathogenic or variants of unknown significance across 110 genes, including 24 founder pathogenic variants.

**Conclusions:** This study provides the first genomic and epidemiological overview of RGDs in the Punjabi population. The findings reveal how genetic, socioeconomic, and cultural factors converge to amplify the RGD burden and highlight the need for affordable molecular diagnostics, inclusive genomic databases, and regional genomic surveillance initiatives in South-Asia.

## Introduction

An RGD occurs intermittently or rarely in the general population. Rarity means only a handful of patients are affected by the disease. The number of estimated RGDs is ∼10,000 (Cipriani et al., 2025), which are caused by both genetic and somatic variations. Characterization of RGDs is challenging due to rarity of incidence and remain mysterious. However, advancing technologies and collaborative efforts of experts are providing novel information to resolve this mystery. In a recent study conducted in 2020, 6172 unique RGDs were identified (Nguengang Wakap et al., 2020). Of these 6172, 69.9% (3,510) had pediatric onset, 11.9% (600) had adult onset, and 18.2% (908) had onset spanning both pediatric and adult groups. Most RGDs occur as natural genetic defects or from missing heritability (Plaiasu et al., 2010, Nguengang Wakap et al., 2020) and are genetically classified as monogenic, polygenic, oligogenic, or chromosomal anomalies (Iourov et al., 2019). RGDs affect approximately 10% of the population (Smith et al., 2022). In some studies, survey point prevalence was expected to be approximately 6.53%, 0.34%, and 0.30% in random populations for common, rare, and ultra-rare genetic diseases, respectively (Fernandez-Marmiesse et al., 2018). RGDs can be grouped into metabolic, neurological, or developmental diseases based on their pathology and phenotype. Diseases that affect fewer than 20 people globally are called ultra-rare diseases (Kar et al., 2024).

The definition of RGDs varies across different geographic regions and healthcare systems (Klimova et al., 2017), but an international definition provided by the World Health Organization (WHO) states that RGD is a medical condition with a specific pattern of signs, symptoms, and clinical findings affecting less than or equal to 1 in 2000 (50/100,000) people in any region (Wainstock and Katz, 2023). We collected information from different global regions about the RGD definition and presented it in Table 1, which shows the differences in RGD definitions across the globe. In Europe, a disease is considered rare when fewer than 50/100,000 people are suffering from it (Dharssi et al., 2017, Haendel et al., 2020). In Latin America, Brazil classifies a disease as RGD when 65/100,000 people are affected, Colombia classifies a disease as rare when 20/100,000 people are affected, and Argentina, Chile, Mexico, Panama, and Uruguay classify a disease as rare when 50/100,000 people are affected (Passos-Bueno et al., 2014, Wainstock and Katz, 2023). In Southeast Asian regions, RGDs are administratively defined as 50, 40, and 10 per 100,000 individuals in China, South Korea, and Taiwan, respectively (Dharssi et al., 2017, Khan, 2023). In South Asian countries, including Bangladesh, India, and Pakistan, no local definitions exist, but they depend on WHO and European definitions.

**Table 1:** Administrative definition of rare diseases and ultra-rare diseases.

| Country | Definition of Rare Disease (Prevalence N/100K)) | Definition of Ultra Rare Disease (Prevalence N/100K) | References |
| --- | --- | --- | --- |
| Argentina | 50 | <2 | (Yu et al., 2021, Richter et al., 2015) |
| Australia | 5-10 | <2 | (Nguengang Wakap et al., 2020, Sharma et al., 2010, Richter et al., 2015) |
| Albania | <50 | <2 | (Perolla et al., 2024) |
| Austria | <50 | <2 | (Moliner and Waligora, 2017) |
| Belgium | <50 | <2 | (Harari, 2016) |
| Bengladesh | 50 | <2 | WHO |
| Bulgaria | <50 | <2 | (Naumova et al., 2022) |
| Brazil | ≤65 | <20 | (Sciascia et al., 2023, Venugopal et al., 2024) |
| Canada | 50 | <2 | (Birkelund, 2019, Sharma et al., 2010) |
| Cyprus | <50 | <2 | (Georgiou et al., 2024) |
| Czech Republic | <50 | <2 | (Richter et al., 2015) |
| Chile | 50 | <2 | (Yu et al., 2021, Richter et al., 2015) |
| China | 50 | - | (WHO)(Lu et al., 2022) |
| Colombia | 20 | <2 | (Yu et al., 2021, Richter et al., 2015) |
| Congo | <50 | <2 | (Lumaka et al., 2018) |
| Croatia | <50 | <2 | (Dumic et al., 2023) |
| Denmark | 20 | <2 | (Birkelund, 2019, Maher et al., 2021, Richter et al., 2015) |
| Ecuador | 10 | <2 | (Zambrano-Mila et al., 2019) |
| Estonia | <50 | <2 | (Tiivoja et al., 2022) |
| Egypt | <20 | <20 | (Chekroun et al., 2025) |
| Ethiopia | 50-100 | <2 | (Mogess and Mihretie, 2024) |
| Europe | <50 | <2 | (Mariz et al., 2016, Sharma et al., 2010) |
| France | <50 | 0.2 | (Harari, 2016, Maher et al., 2021, Richter et al., 2015) |
| Finland | <50 | <2 | (Maher et al., 2021) |
| Germany | <50 | 0.2 | (Moliner and Waligora, 2017, Richter et al., 2015)<br>https://www.narse.de/en/translate-to-englisch-ueber-das-narse |
| Georgia | <50 | <2 | (Czech et al., 2020) |
| Ghana | No official definition | <2 | https://frontlinegenomics.com/world-of-genomics-ghana/ |
| Greece | <50 | <2 | (Maher et al., 2021) |
| Hong Kong | <5 | <2 | (Chung et al., 2022) |
| Hungry | <50 | <2 | (Sciascia et al., 2023) |
| India | 50 | <2 | (WHO) |
| Iceland | <50 | <2 | (Jensson et al., 2023) |
| Indonesia | No official definition | <2 | (Sihombing et al., 2021) |
| Ireland | <50 | <2 | (Modell et al., 2016, Richter et al., 2015) |
| Iran | 20 |  | (Birkelund, 2019) |
| Italy | 50 | <2 | (Maher et al., 2021, Richter et al., 2015, Sciascia et al., 2023) |
| Japan | 40 | <2 | (Nguengang Wakap et al., 2020, Sharma et al., 2010, Maher et al., 2021)<br>(Mariz et al., 2016, Perolla et al., 2024) |
| Jordan | <50 | <2 | (Obeidat et al., 2024) |
| Kenya | No official definition | <2 | https://rarediseasekenya.org/resources/ |
| Kazakhstan | 5 | <2 | (Aymé, 2012) |
| Latin America | <65 | <2 | (Baldovino et al., 2025) |
| Lithuania | <50 | <2 | (Cerkauskaite-Kerpauskiene et al., 2025) |
| Luxembourg | <50 | <2 | https://www.iml.lu/sites/default/files/2021-08/orphan-drugs.pdf |
| Mali | No local definition exists | <2 | (Landoure et al., 2016) |
| Malaysia | <25 |  | (Shafie et al., 2020) |
| Mexico | ≤50 | <2 | (Richter et al., 2015, Sciascia et al., 2023) |
| Morocco | No official definition | <2 | (Charoute et al., 2014) |
| New Zealand | No national consensus | <2 | (Theadom et al., 2019) |
| Nigeria | No official definition | <2 | (Adeyemo et al., 2018) |
| Netherlands | <0.67 |  | (Richter et al., 2015, Czech et al., 2020) |
| Norway | 50 | <2 | (Lorentsen et al., 2025) |
| Pakistan | 50 | <2 | (WHO) |
| Panama | 50 | <2 | (Sciascia et al., 2023) |
| Poland | <50 | <2 | (Domaradzki and Walkowiak, 2023, Maher et al., 2021, Richter et al., 2015) |
| Peru | 1 | <2 | (Bazalar-Montoya et al., 2024) |
| Philippines | 5 | <2 | (Sciascia et al., 2023) |
| Portugal | <50 | <2 | (Marta et al., 2024) |
| Qatar | <50 | <2 | (Chekroun et al., 2025, Elfatih et al., 2024) |
| Romania | <50 | <2 | (Drugan et al., 2002) |
| Russia | 10 | <2 | (Yu et al., 2021, Richter et al., 2015) |
| Saudi Arabia | <50 | <2 | (Alqahtani et al., 2023) |
| Singapore | <5 | <2 | (Chung et al., 2022) |
| South Africa | <50 | <1 | (Richter et al., 2015) |
| South Korea | 40 | <2 | (Maher et al., 2021, Song et al., 2012) |
| Spain | <50 | <2 | (Sciascia et al., 2023, Richter et al., 2015) |
| Sri Lanka | <50 | <2 | (Sirisena and Dissanayake, 2019) |
| Sweden | 10-50 | <2 | (Richter et al., 2015) |
| Switzerland | 50 | <2 | (Wehrli et al., 2024) |
| Thailand | <5 | <2 | (Chung et al., 2022) |
| Taiwan | 10-100 | <2 | (Sharma et al., 2010, Maher et al., 2021, Richter et al., 2015, Nguengang Wakap et al., 2020) |
| Turkey | 50 | <2 | (Sciascia et al., 2023, Richter et al., 2015) |
| United Kingdom | <50 | <2 | (Gentilini et al., 2025, Maher et al., 2021) |
| United Arab Emirates | <50 | <2 | (Bizzari et al., 2023) |
| Ukraine | <50 | <2 | (Boyarchuk et al., 2024) |
| United States of America | <60 | <2 | (Sciascia et al., 2023, Sharma et al., 2010, Maher et al., 2021) |
| Uruguay | 50 | <2 | (Passos-Bueno et al., 2014, Wainstock and Katz, 2023) |
| Vietnam | <50 |  | (Nguyen et al., 2025) |
| Zimbabwe | 50 | <2 | (WHO) |

The definitions of "rare" and "ultra-rare" diseases vary across countries and regions, primarily based on disease prevalence within the population, as shown in Table 1. RGDs are generally defined based on their low prevalence within a population, with thresholds varying by country. Ultra-rare diseases are typically considered to affect fewer than 1 in 50,000 individuals, although official definitions are less common. Hyper-rare diseases are likely to exist among undiagnosed RGD patients. The major challenges include the characterization of these disease genotypes and phenotypes, given that for most of these diseases, only a single patient will be diagnosed (Landrum et al., 2020). Pakistan currently lacks an official definition, but international standards are often used as references.

From a medical perspective, the characterization of an RGD depends on the broader diversity of diseases and symptoms that can vary from disease to disease as well as within the same disease (Thevenon et al., 2016). The same disease can have variations in clinical manifestations from person to person, which are differentiated into subtypes of the same disease, and it remains challenging in the diagnostic journey due to variable factors (Frederiksen et al., 2022). Furthermore, each RGD has a different effect on life expectancy: some are fatal at birth, some are degenerative and life threatening, whereas others are compatible with a normal life if diagnosed in time and properly managed and/or treated (Rode, 2021).

Due to the wide range of diversity and complexity, it is difficult to properly recognize an RGD at earlier stages, resulting in a longer diagnostic odyssey (Janku et al., 1980, Cooper et al., 2013, Ahluwalia et al., 2009). Generally, the first line of diagnosis is a clinical diagnosis, including routine hematology and biochemistry tests and radiographic examinations. Once a provisional diagnosis is established, confirmatory diagnostic tests are necessary to make a definitive diagnosis. Classically, biochemistry and histopathology practices have been used to identify pathology at the protein level. Recently, karyotyping and in situ hybridization methods have been applied on a vast scale to identify pathologies at the genetic level. These methods, together with polymerase chain reaction-based assays, have been widely accepted for their target-oriented accuracy (Hartley et al., 2020). However, the etiology of many RGDs has not yet been established. This unknown etiology has led to the use of advanced genomic techniques for RGD diagnosis. Despite the known variants, there are unknown variants that probably have 10-90% pathogenic effects (Richards et al., 2015). The growing chances of ambiguity in the results of RGD are due to these unknown variants called variants of uncertain significance (VUS).

Recently, short- and long-read NGS (Merker et al., 2018, Mizuguchi et al., 2019) and single-cell genomic technologies (Katsanis and Katsanis, 2013) have been used for RGD diagnosis. NGS has accelerated the precise diagnosis of RGDs with a confirmed outcome of 25-50% (Li et al., 2018). Comparative analysis of clinical data and NGS data, together with advanced bioinformatics methods, has proven to be a powerful technique for accurate diagnosis (Wenger et al., 2017). Three types of NGS approaches have been applied: whole-genome sequencing (WGS), whole-exome sequencing (WES), and targeted capture sequencing. These approaches can be used for both variant discovery and copy number variations (Akhtar et al., 2022). WES is a leading diagnostic strategy (Masri et al., 2022); however, one-third of RGDs remain undiagnosed because of technical limitations in determining variations in non-coding regulatory genomic regions (Frésard and Montgomery, 2018). To overcome these limitations of WES, WGS can identify non-coding region variants and is able to identify 99% of the significant risk of RGD (Adhikari et al., 2020, Woerner et al., 2021) in addition to those possible with WES (Wang et al., 2015, 100, 2021, Manickam et al., 2021, Malinowski et al., 2020, Lavelle et al., 2022). However, in cases when a provisional diagnosis is strong and patient had a family history of a particular disease, it is possible to utilize targeted capture sequencing instead of WGS and WES. In addition to these conventional NGS applications, single-cell genomic technologies have the potential to identify pathologies at the cellular level. Single-cell technology defines cellular heterogeneity at the cell level; however, specialized computational tools are required for comparison with disease phenotypes (Auerbach et al., 2021).

Despite the availability of these advanced technologies, RGD diagnosis remains a challenge because these technologies have not been optimized for this purpose owing to limited testing options. Challenges in RGD diagnosis also lead to challenges in treatment. Most RGDs involve neurodevelopmental and metabolic pathologies (Tarailo-Graovac et al., 2016). Affected patients are on a life-death race, and death tolls always remain higher. Higher mortality and limited lifespans have made it challenging to devise treatments using conventional methods (Black et al., 2015, Carmichael et al., 2015). Most RGDs do not have a treatment, but different therapeutic regimens are used only to manage the clinical presentation (Clark et al., 2019, Farnaes et al., 2018, Owen et al., 2021, Sanford et al., 2019, Willig et al., 2015). Due to the rarity of RGDs, drug development is challenging, and medicinal treatment exists for only 5% of RGDs to date (Nguengang Wakap et al., 2020). However, recent advances in genomics and drug development have opened new avenues for discovery. Moreover, biobanking, organ-on-chip devices, and *in silico* docking technologies have the potential to validate novel drugs (Sequeira et al., 2021).

It is worth noting that among the general population, six to eight people out of a hundred are carriers of RGDs. There is no significance of these carrier individuals, but if two individuals marry and carry the same genetic abnormality, there is a 25% chance of inheriting a genetic abnormality per pregnancy. It can be frequent to “be affected by” an RGD in a family carrying RGD but it is “rare” to find a family carrying RGD (Rode, 2021). Marrying a cousin (consanguinity) in such families can lead to a higher incidence of RGD. The consanguinity rate varies among populations based on religion, culture, and politics. The global distribution of consanguinity is 10.4% of the entire population (Bittles and Black, 2010). Consanguineous marriages are prevalent in North Africa, West, and South Asia. The consanguinity rates of some countries in this region are reported as Saudi Arabia, 30% (Warsy et al., 2014), Libya, 38% (Abudejaja et al., 1987), Qatar, 54% (Bener and Alali, 2006), and Pakistan over 80% (Akhtar et al., 2019, Small et al., 2017). The consanguinity level is comparatively low (1-1.5%) in European, South American, and Australian populations depending on the social demographic effect (Liascovich et al., 2000, McCullough and O’Rourke, 1986).

Numerous barriers to RGD patients and carriers are observed in developing countries. Pakistan is one such country where RGD patients suffer from socioeconomic crises. In addition, the Pakistani population has a consanguinity rate of more than 80%. This higher consanguinity at the population scale increases the risk of genetic diseases, including RGD (Akhtar et al., 2019, Aslamkhan et al., 1969, Zar et al., 2020, Wasim et al., 2023, Aslamkhan et al., 2023). A high consanguinity rate leads to reproductive loss, risk of abortion, and neonatal or postnatal death (Romdhane et al., 2019). However, no statistics exist on the incidence of RGDs in the Pakistani population. It is important to look for existing RGDs, causal genes, pathogenic variants, and their relationships with demographic factors in the Pakistani population (Akthar, 2019). The Pakistani population is generally comprised of Caucasian ethnic groups (Mohyuddin et al., 2002). The major ethnic groups include Punjabi, Pathan, Sindhi, Saraiki, Balochi, Kashmiri, and Kalash. Punjabi people comprise the largest group, with approximately 150 million people. Punjabi people not only live in Pakistan but also have major populations in India (∼37 million), Canada (∼1 million), United States (∼0.25 million), and England (∼0.75 million).

All these Punjabi populations were homed in the Punjab state of British India until 1947, when it was divided into two countries, India and Pakistan. Following this partition, many Punjabis moved out of India due to religious, political, or identity crises. Many of these migrating populations moved to the United States, Canada, England, and other European countries only one or two generations ago and thus share the same genetic backgrounds as the Pakistani and Indian Punjabi populations. In Pakistani Punjab, the majority of Punjabi people practice Islam and are practicing Muslims, while in India, the majority of Punjabi follow Sikhism. Sikhism prohibits consanguinity up to seven generations, but it is common in Pakistani Punjab. This is the reason why Pakistani Punjabi have consanguinity rates of over 80% (Akhtar et al., 2019), while in Indian Punjab, it is less than 5% (Acharya and Sahoo, 2021). This higher consanguinity is not only common among Pakistani Punjabis but also in the Muslim world in general (Ghanim et al., 2023). It is not only religion but also culture, family wealth, power politics, and other associated factors that have historically led to increased consanguinity practices.

In this study, we reported a recently built RGD provincial registry in the Punjab region of Pakistan. We registered the maximum possible number of families of Punjabi origin with confirmed RGD diagnosis and obtained information on RGD, causal gene, pathogenic variant, consanguinity, and other demographic factors. Compiling all this data, we presented the incidence of RGDs, causal genes, pathogenic variants, and the role of consanguinity during 2021-2023 in this manuscript.

## Methods Study Settings

The study was conducted from January 2021 to December 2023 at the PTGDPRI at Sir Ganga Ram Hospital, Lahore, Pakistan and National Center for Excellence in Molecular Biology (CEMB), University of Punjab (PU), Lahore, Pakistan. An informed consent was obtained from all registered families and the study was approved by institutional review board of CEMB under letter no. IRB-5/24.

### Sampling Technique

The snowball sampling technique was used to include the maximum possible RGDs in the registry presented in this study. The PTGDPRI offers chorionic villus sampling (CVS) for prenatal screening of genetic diseases; thus, RGD carrier families walked in for CVS have been registered in this registry since January 2021 after informed consent.

### Inclusion and exclusion criteria

The inclusion criterion for our study (also the registry) was any family with a confirmed RGD diagnosis. All diagnoses were based on WGS, WES, or targeted capture sequencing. In case a patient was suspected of having an RGD, the patient was followed up until a final diagnosis was made. Patients with suspected disease for whom a final diagnosis was not made by December 31, 2023, were not included or presented in this study.

### Patient Data Collection

Patient data were recorded using a registry questionnaire, including demographic information such as ethnicity, caste, district, consanguinity, family income, and total number of children in the family, including affected, died and normal children. In addition, gravida (G [total pregnancies]), parity (P [viable births]), and abortions (A), collectively termed GPA information, were also recorded.

### Genetic data

RGD genotypes were registered for 91 families in the current registry. This genotype information was obtained from trio massive parallel NGS (WGS, WES, or targeted capture sequencing) of the proband and both parents. WGS was performed either by referring clinician’s office or a third party, and only genotypes of potential clinical interest, including pathogenic, likely pathogenic, or variants of unknown significance, were registered.

### Healthy Controls Data

Consanguinity data of 1011 healthy couples without RGDs were obtained from the Punjab Consanguinity Survey (Shenk et al., 2024). These data were used to compare the RGD carrier families with healthy couples.

### Consanguinity Analysis

We conducted a descriptive comparison of consanguinity patterns in RGDs. Data were collected from demographic sources based on their link to consanguineous marriages. We report the frequencies of the overall rates of cousin marriages. This analysis provided a more precise breakdown of the proportion of marriages between first cousins, second cousins, relatives, and unrelated individuals among RGD carrier families. The consanguinity information of RGD carrier families and healthy couples from Shenk et al. (2024) (Shenk et al., 2024) was tested for the association of consanguinity with RGDs using the chi-square test.

### Variant annotation

After recording each patient/proband and RGD carrier family in the registry, primary, secondary, and variant of unknown significance information were also recorded in the registry. All variants were annotated against their registered single nucleotide polymorphism (SNP) identification numbers (rsIDs) allocated by the database of SNPs (dbSNP) (Sherry et al., 1999) or nucleotide identification (NM) allocated by the ClinVar database. The allele frequency of each registered variant in our registry was calculated using the following formula:

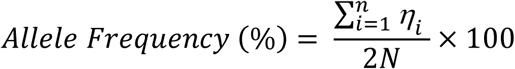

Where 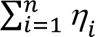 is the sum of alleles of carriers for each variant and 2N is the sum of ploidy (total chromosomes) in the registry.

The allele frequency of each registered variant worldwide was annotated using rsIDs from the gnomAD v4.0 database to understand the global distribution of variants and identify population-specific variants.

### Statistical Analysis

We performed descriptive statistics, including frequencies and percentages, for qualitative variables. Central tendency and distributions were measured using the mean and standard deviation. Crosstabs were constructed between the variables to establish relationships. Chi-square was applied to determine association between qualitative variables with p-value threshold of 0.05. SPSS v22.0, Microsoft Excel 2016, GraphPad Prism 10 or R Studio (2025.05.1+513) packages *tidyr* and *ggplot2* were used for these analyses and visualizations.

## Results

### Identification and incidence of RGDs

The PTGDPRI is the only public institute dedicated to RGDs in Punjab and Pakistan. This institute is run by the provincial government of Punjab and has maintained an RGD registry since January 2021. We have presented data spanning 2021-2023 in this study. We registered patients visiting the PTGDPRI for clinical services, including diagnosis, prenatal screening, or genetic counselling. Although this institute is a provincial body, patients from other geographic regions and provinces of the country also visit for clinical services (Supplementary Figure 1A). RGDs are broadly distributed across many districts, but most RGD-city combinations involve only a single affected family. However, certain cities, such as Lahore, Faisalabad and Rawalpindi, which are metropolitan, show clusters of higher frequencies for specific RGDs, with up to six cases observed in some instances. These data underscore the importance of region-wide genetic screening, as RGDs are present throughout the country (Supplementary Figure 1B).

In our data spanning 36 months from January 2021 to December 2023, 167 families visited the PTGDPRI. These families presented 72 RGDs of pediatric onset as primary diagnosis. A list of these 72 RGDs along with their incidence within the current registry is provided in Supplementary Table 1. These 167 families represented 118 probands and 391 children. Of the 391 children, 293 were affected by RGDs and 188 had died. Of these 188 children who died, 70 died without any diagnosis, including both pre-and post-mortem investigations.

All 167 families were aware of the incidence of RGD in the family and underwent CVS for an ongoing pregnancy. However, only 91 families submitted detailed genetic data at the time of registration, while others only submitted a clinical final diagnosis (confirmed on genetic testing) with a referral registration form submitted by the referring clinician’s office. All 91 families who submitted genetic data were tested using massive parallel NGS for short and structural variants. Short variant testing was performed for 56 families, and 35 families were tested for structural variants in addition to short variant testing. Eight of these families underwent whole-genome sequencing (WGS), 56 underwent whole-exome sequencing (WES), and 27 underwent targeted capture sequencing. This information is presented in Table 2. In 91 cases, 72 RGDs were associated with primary variants whereas other 34 RGDs were associated with secondary variants (Supplementary Table 2A). All these RGDs showed an autosomal recessive inheritance pattern, except for adrenoleukodystrophy, G6PD deficiency, hemophilia, and mental retardation type 5, which are known to be X-linked recessive. These RGDs were caused by pathogenic, likely pathogenic or VUS in 110 genes (Supplementary Table 2(B &C)). A primary variant was identified in all 91 cases, with 58 secondary variants and 31 variants of unknown significance (Supplementary Table 2D). All these variants were SNVs except one CNV in TJP2 gene. A summary of the pathogenic classification of these variants is provided in Table 3.

**Table 2:** The number of Diagnoses by different genomic techniques.

| **Number of Diagnoses made by Short and Structural Variant Discovery Methods** |  |  |
| --- | --- | --- |
|  | Short Variant Discovery | Structural / Copy Number Variation |
| Short Variant Discovery | 91 | 35 |
| Copy Number Variation | 35 | 35 |
| **Sequencing Techniques Utilized for Diagnosis** |  |  |
| Whole Genome Sequencing | Whole Exome Sequencing | Targeted Capture Sequencing |
| 8 | 56 | 27 |

**Table 3:** Classification of variants identified in patients in current study.

| **Variant Class** | **Primary**<br>**Variants** | **Secondary**<br>**Variants** | **Variants Of Unknown**<br>**Significance** |
| --- | --- | --- | --- |
| Frameshift Likely Pathogenic | 1 | 5 | 0 |
| Frameshift Pathogenic | 6 | 0 | 0 |
| In Frame Pathogenic | 1 | 0 | 0 |
| In Frame Uncertain Significance | 2 | 1 | 3 |
| Large Inversion | 1 | 0 | 0 |
| Likely Pathogenic | 5 | 11 | 0 |
| Loss Like Pathogenic | 2 | 0 | 0 |
| Loss Pathogenic | 1 | 0 | 0 |
| Missense Pathogenic | 18 | 18 | 0 |
| Missense Likely Pathogenic | 14 | 6 | 0 |
| Missense Uncertain Significance | 13 | 7 | 20 |
| Nonsense Pathogenic | 12 | 1 | 0 |
| Nonsense Likely Pathogenic | 5 | 2 | 0 |
| Pathogenic | 1 | 1 | 0 |
| Silent Pathogenic | 0 | 1 | 0 |
| Silent Uncertain Significance | 1 | 0 | 1 |
| Splicing Likely Pathogenic | 1 | 0 | 0 |
| Splicing Pathogenic | 3 | 2 | 0 |
| Splicing Uncertain Significance | 1 | 0 | 1 |
| Vus | 3 | 3 | 6 |
| Total | 91 | 58 | 31 |

Of these 72 RGDs, 42 were of metabolic origin, including 37 inborn errors of metabolism; 11 neurodevelopmental diseases; five immunodeficiency diseases; three congenital, endocrine or connective tissue diseases; and one lymphatic, skeletal, hematological, or bone disease. In our registry, we had 21 families with, 20 with PFIC [including PFIC I, PFIC II, PFIC IV, and PFIC V], 15 with MPS [including MPS I, MPS IIIA, MPS IIIB, and MPS IV], seven with Gaucher disease, five with methyl malonic aciduria or cystic fibrosis, four with glycogen storage disease, hemophilia, or Sandhoff disease, three with X-linked adrenoleukodystrophy, severe combined immunodeficiency (SCID), osteoporosis, propionic acidemia, gangliosidosis, or citrullinemia, two with congenital adrenal hyperplasia, congenital hydrocephalus type 3, glutaric aciduria, Pompe disease, spinal muscular atrophy, Tay Sach disease, or tyrosinemia. For the remaining RGDs in the registry, only a single family was registered per RGD. The number of RGD carrier families is shown in Figure 1. In our registry, 36 of the 72 RGDs were categorized as ultra-rare diseases according to the global RGD directories (Supplementary Figure 2). No hyper-rare disease was found in the current cohort. Two RGDs, PFIC and gangliosidosis, known as ultra-rare diseases globally, were not rare in our registry, with 20 and three carrier families, respectively. Also, we reported gangliosidosis and its subtypes (Sandhoff disease and β-galactosidase deficiency) separately based on their differing genotypes and phenotypes. In addition, the five most common diseases, NP, PFIC, MPS, Gaucher disease, and methyl malonic aciduria, contributed to approximately 40% of the burden in this registry.

**Figure 1:**
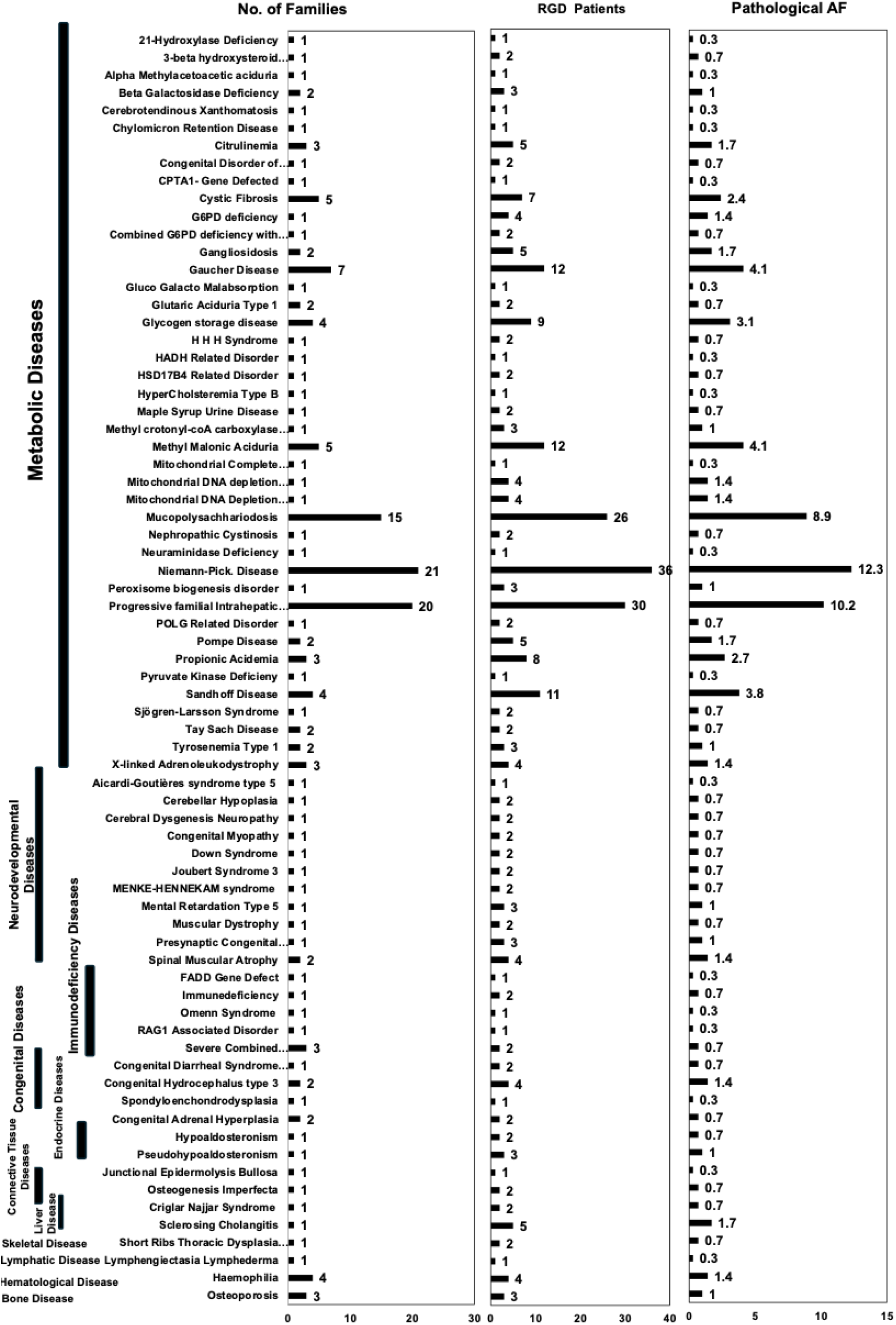
Distribution of RGDs among studied families. Left panel shows the number of families affected by each RGD, highlighting the variable incidence across diseases. Middle panel presents the incidence of RGDs within the cohort, reflecting the overall disease burden in the population. Right panel displays the pathological allele frequency within, indicating compound burden of RGD-causing allele and providing insights into the genetically distribution across diseases.

Of the 91 primary variants, 76 were single nucleotide variants (SNVs), six were deletions, five were insertions, three were CNVs, and one was an inversion. Among the 58 secondary variants, 45 were SNVs, 11 were deletions, one was an insertion, and one was a substitution.

The current registry is naïve and is still recruiting patients in collaboration with clinicians, hospitals, and PTGDPRI’s province-wide district centers. To date, a snowball sampling strategy has been used, and our registry is not representative of population-wide RGD statistics. However, this is the first study to identify the incidence of many RGDs in the region. Thus, we did not use any population-wide statistics to report RGD incidence but the number of patients and pathological allele frequencies (AF) within the registry. We have shown the number of patients per RGD in Figure 1, which illustrates that NP has the highest incidence in our registry, followed by PFIC and then MPS. This incidence is representative of the accumulated burden of affected children with each disease. We calculated the pathological allele frequency of each registered RGD within our registry, as shown in Figure 1. We counted the zygosity of RGD carriers or affected children in this registry and calculated the pathological allele frequency for each RGD. The allele frequencies in our dataset ranged from 0.34% to 5.80% in the registry. The highest observed pathogenic alleles were observed in MPS (5.8%), PFIC (5.46%), and NP (3.75%) in our registry.

### Epidemiological profiling of RGD carrier families

In total, 167 families were included in our dataset, with 293 affected children. We extended our search criteria and counted the total number of children, including probands, died, and unaffected children. On average, each family had 2.34±1.1 children per family. Of these, 0.71±0.6 were diagnosed with RGD. These families also reported that their 1.13±1.1 children died due to an RGD. Ultimately, these families had only 0.59±0.7 normal children. The total number of children per couple (2.34±1.1) in this registry was lower than the national average of 3.6 per couple (Shenk et al., 2024), suggesting lower fertility in RGD carrier families. Of the 391 children, 188 died, with a mortality rate of 480 children per 1000, which is significantly higher than the national child mortality rate (62/1000) (Shenk et al., 2024). These statistics of higher mortality and fewer normal children in RGD carrier families are indicators of poor quality of family life. The burden of RGD per family (0-1) was calculated as the ratio between affected and total children in each RGD family. The overall burden of RGD children per family is presented in Figure 2A. Mean burden of affected children among all families was 0.81±0.24. The maximum RGD burden observed was 1.0 in 95 families, which means that all born children were affected by an RGD. However, the lowest disease burden was 0.3, observed in six families with a history of RGD carriers who walked into PTGDPRI for prenatal screening and genetic counselling.

**Figure 2:**
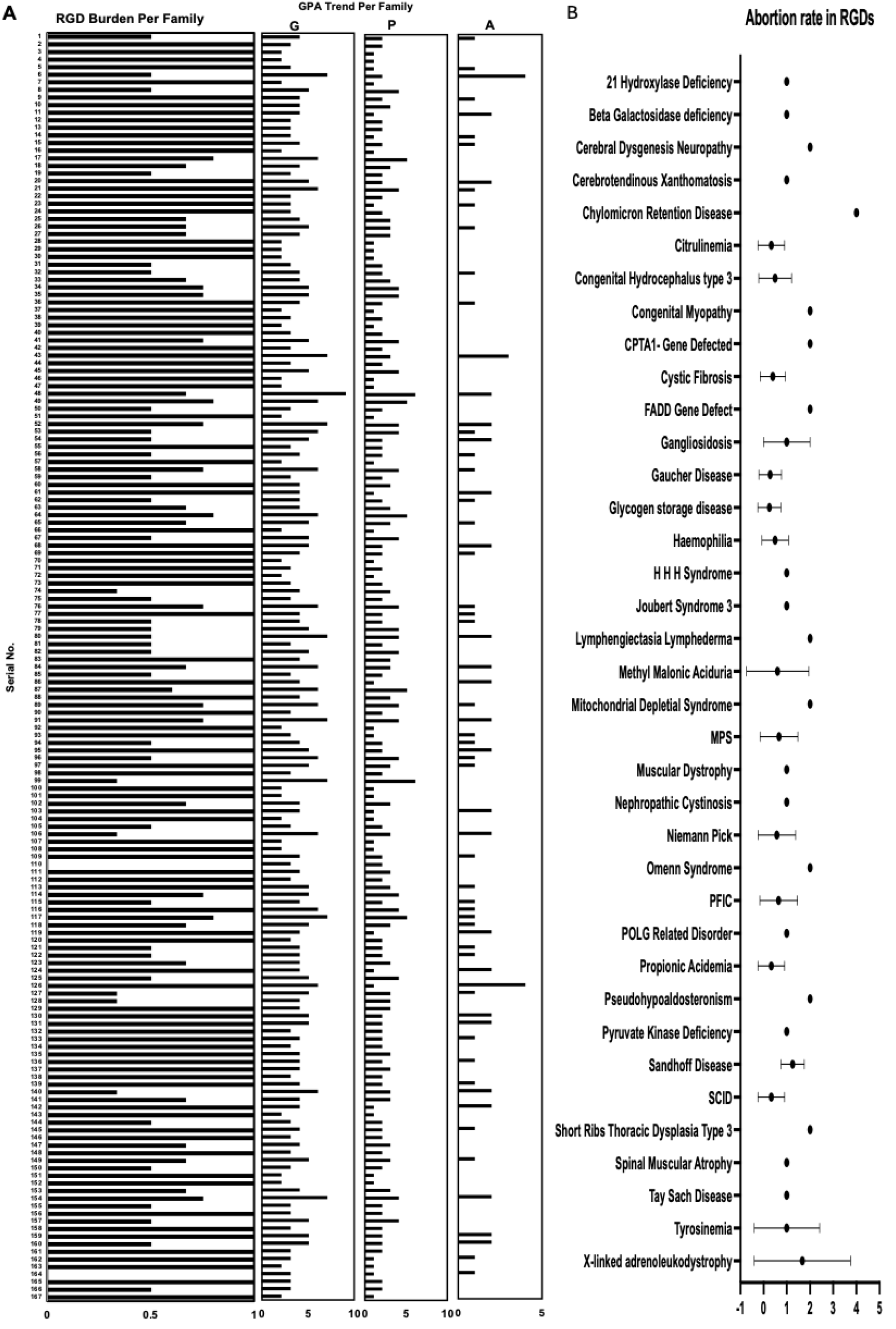
GPA profile and abortion rate among RGDs families. A: Family wise GPA profile among RGDs families is shown. This analysis highlights the variations in reproductive history and pregnancy outcomes within affected families B: Distribution of abortion rate by RGDs is shown indicating the association between specific RGDs and increased risk of abortion

The family wise plot of GPA revealed important reproductive health patterns (Figure 2A). The G per family generally ranged between one and nine (mean 3.92±1.4), with P between one and six (mean 2.34±1.1), and A between one and four (mean 0.58±0.8). Most of these families were registered while they were pregnant, and the outcome as P or A was not yet understood. The close observation of P with G suggests consistently lower P underscoring the impact of A. These findings highlight the general reproductive trends in the cohort, with high desired fertility rates (G) and moderate outcomes as viable births (P) due to the influence of A. Such insights are valuable for maternal health interventions and understanding reproductive patterns in RGD carrier families. These important findings suggest that the desired fertility rates are higher, but the actual fertility rates are lower among RGD carrier families.

We also identified 37 RGDs associated with the incidence of abortion in our registry (Figure 2B). These 37 RGDs were observed in 66 families. Moreover, we observed that couples who are carriers of chylomicron retention disease or X-linked adrenoleukodystrophy are at a higher risk of abortion. Although we only had one family with chylomicron retention disease, this family had a history of four abortions. Chylomicron retention disease is a life-threatening disease caused by a rare autosomal mutation in the SAR1B gene, and patients cannot absorb fats or fat-soluble vitamins due to the lack of chylomicron synthesis and secretion. This is family 6, as shown in Figure 2A, had a G:P:A of 7:2:4, which included one current pregnancy with four abortions, and only two fetuses survived. Among these two, only one was normal, and the other was a patient with chylomicron retention disease.

In addition to chylomicron retention disease, X-linked adrenoleukodystrophy also showed the highest abortion rate. Three families with X-linked adrenoleukodystrophy were included in our registry (families 125, 126, and 127 in Figure 2A). These families had G:P:A ratios of 5:4:0, 6:1:4, and 5:3:1. Family 125 did not have any abortions, but family 126 had the highest abortion incidence of four fetuses. Family 127 had six pregnancies, including the current pregnancy, but only one fetus survived and was diagnosed with X-linked adrenoleukodystrophy.

### RGD burden across ethnic groups

Although our institute is based in Punjab, RGD patients of both Punjabi and non-Punjabi ethnic origins are part of our registry. These non-Punjabi ethnic groups are either residents of the province of Punjab or registered with the PTGDPRI for clinical services from other provinces. According to ethnicity, there were 118 RGD families of Punjabi ethnic origin, 34 Pathan, five Saraiki, four Sindhi, three Kashmiri, two Balochi, and one Hindko. Our data show clear patterns of differential distribution of RGDs in ethnic groups that emphasize the importance of inclusion in RGD studies. These trends were most significant when comparing the Punjabi population with all other populations, as demonstrated in Figure 3. The Punjabi population demonstrated a significantly higher incidence of RGDs than the non-Punjabi population. A close examination of Figure 3 shows that 58 RGDs are present in the Punjabi population in our registry. Of these 58 RGDs, 47 were specific to the Punjabi population in the current registry, indicating a possible founder effect or a higher prevalence of pathogenic alleles within the population.

**Figure 3:**
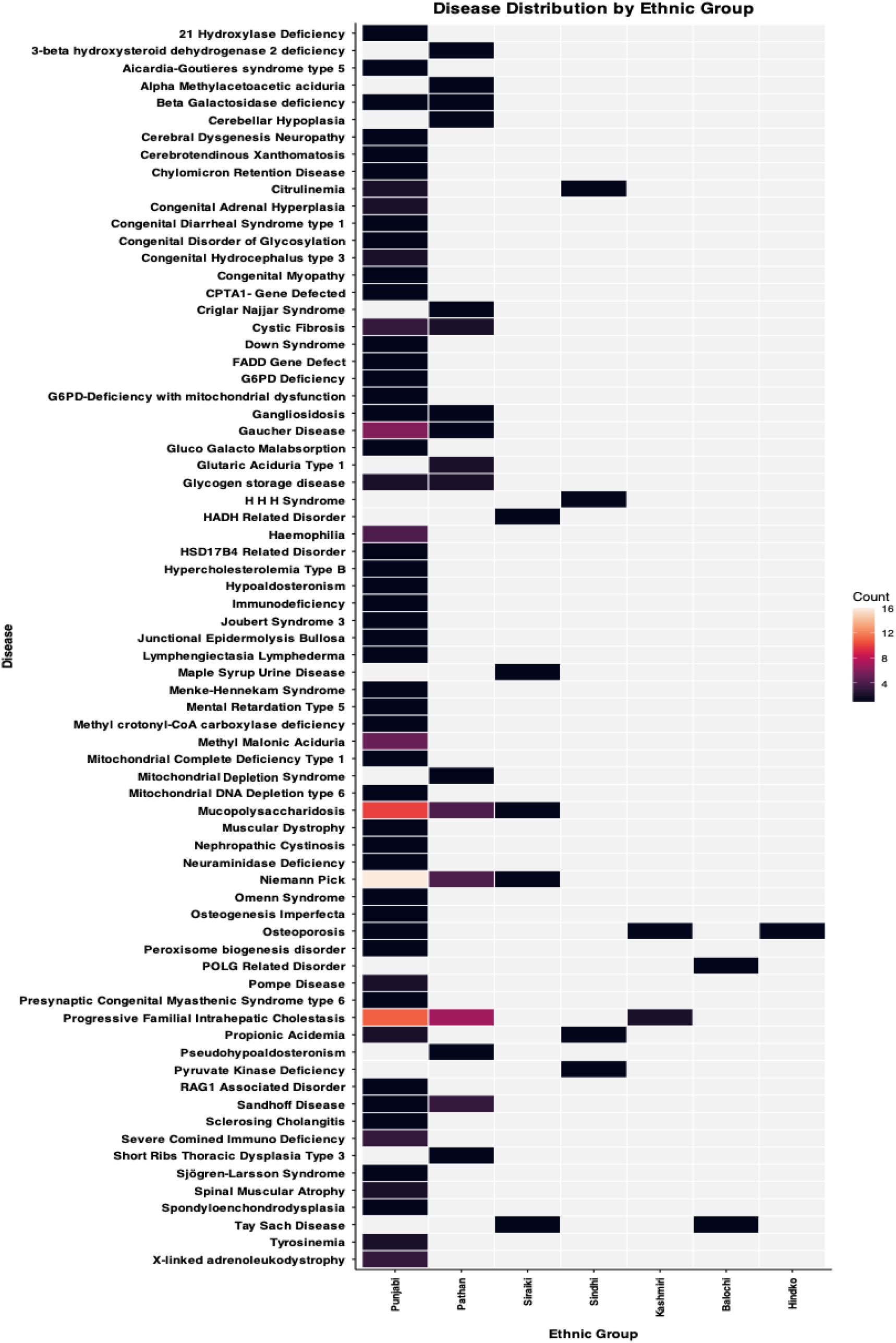
RGDs distribution among ethnic groups. RGDs are distributed among different ethnic groups, highlighting differences in the frequency of RGD within each group

While looking into the non-Punjabi population, we found 16 RGDs present in the Pathan population. Of these 16 RGDs, eight were present only in the Pathan population. These eight RGDs include 3-β hydroxysteroid dehydrogenase 2 deficiency, α-methyl acetoacetic aciduria, cerebellar hypoplasia, Crigler-Najjar syndrome, glutaric aciduria type I, mitochondrial depletion syndrome, pseudohypoaldosteronism, and short rib thoracic dysplasia type 2. Five RGDs, including cystic fibrosis, gangliosidosis, Gaucher disease, glycogen storage disease, and Sandhoff disease, were cumulatively present in the Punjabi and Pathan populations. The other three diseases (NP, MPS, and PFIC) were shared by more than two ethnic groups.

Five RGDs were present in the Saraiki population. Of these five, two RGDs (HADH-related disorder and Maple Syrup Urine Disease) were only present in Saraiki ethnic families. Four RGDs were observed in the Sindhi population. Among these RGDs, two (HHH Syndrome and Pyruvate Kinase Deficiency) were specific to the Sindhi population in this registry. Moreover, two other RGDs (citrullinemia and propionic acidemia) were shared between the Punjabi and Sindhi populations. Among the two Balochi ethnic families, the POLG-related disorder was observed in one family and was specific to the Balochi population. However, Tay-Sachs disease, the second RGD observed in the Balochi ethnic group, was also found in the Saraiki population, consistent with the colocalization of both groups in the Southern Punjab region. In the Hindko ethnic group, we observed only one disease (osteoporosis), which was also shared among the Punjabi and Kashmiri ethnic groups, consistent with the historical closer geographic existence of the Hindko, Kashmiri, and Punjabi populations.

### RGD distribution across castes

Ethnicity was further subdivided into castes. The number of families carrying an RGD within each caste is shown in Figure 4. In Punjabi castes, highest RGD incidence was recorded in Arain (22, 13.2 %) and Rajput (22, 13.2%), followed by Sheikh (14, 8.4 %), Butt (9, 5.4%), and Jutt (9, 5.4%) castes. On a disease-wise pattern, we found that 21 RGDs were distributed across the castes and 51 RGDs were specific to only one caste, while others, including MPS, NP, osteoporosis, and PFIC, were distributed across the castes. Among Rajput caste, seven RGDs were caste-specific including Cerebrotendinous Xanthomatosis, congenital diarrheal syndrome, HSD17B4-related disorder, hypercholesterolemia type B, Menke-Hennekam Syndrome, RAG1-associated disorder and Sjögren-Larsson Syndrome. Six RGDs were specific to the Arain caste, including congenital disorder of glycosylation, CPTA1-gene defect, junctional epidermolysis bullosa, mental retardation type 5, neuraminidase deficiency, and Omenn syndrome. Five RGDs were specific to Sheikh caste, including cerebral dysgenesis neuropathy, FADD gene defect, osteogenesis imperfecta, spinal muscular atrophy, and tyrosinemia. The Jutt caste had three caste-specific RGDs, including hypoaldosteronism, mitochondrial complete deficiency, and presynaptic congenital myasthenic syndrome. Among the Butt caste, two RGDs (Joubert syndrome and peroxisome biogenesis disorder) were caste-specific.

**Figure 4:**
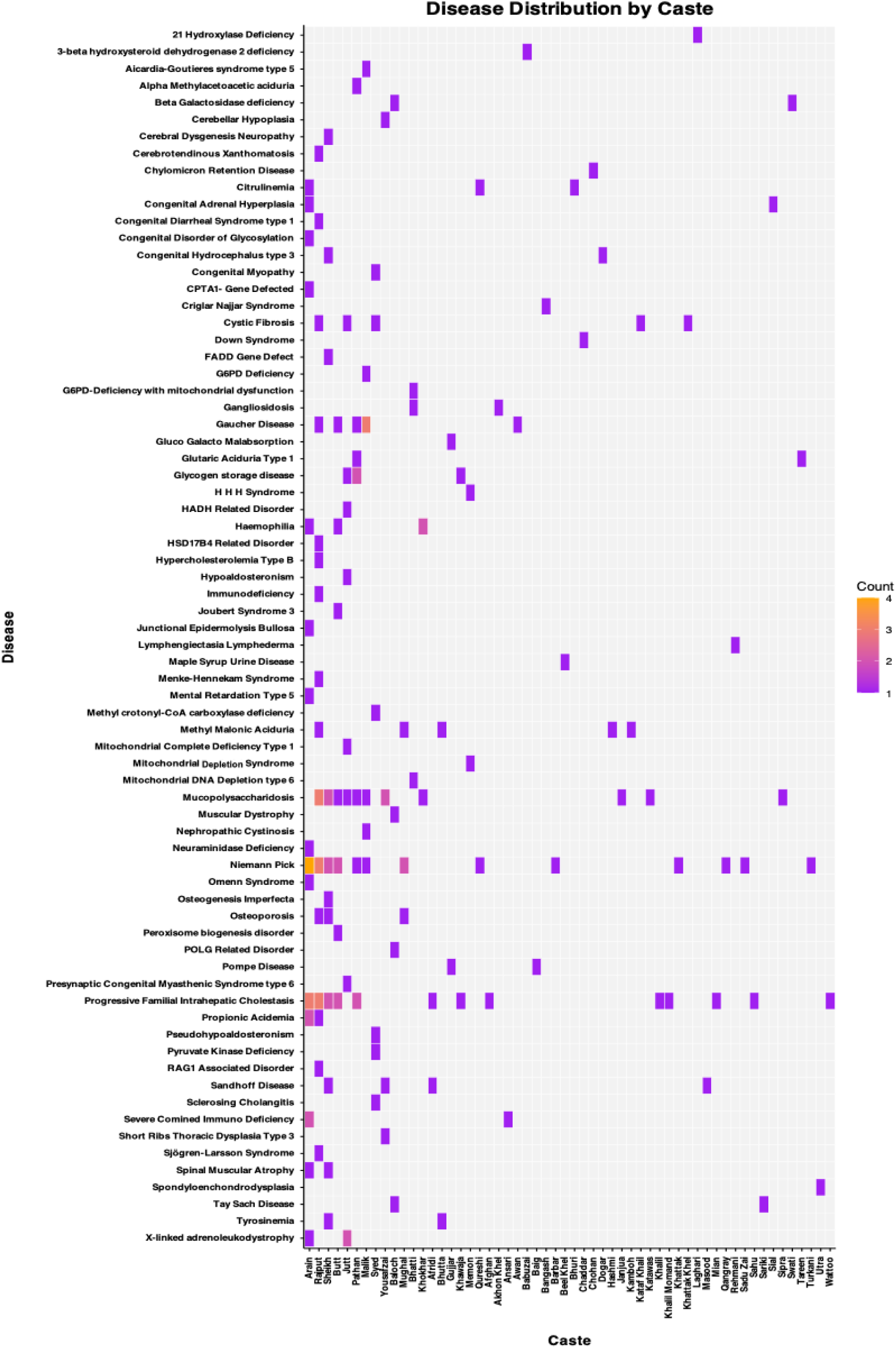
RGDs distribution among castes. Distribution of RGDs by castes is shown. It demonstrates how the occurrence of specific disease varies among castes reflecting underlying genetic patterns

Looking deeper into caste-specific disease incidence, we found that most castes had a specific RGD incidence, except Arain, Baloch, Barber, Bhatti, Butt, Gujjar, Jutt, Khawaja, Khokhar, Malik, Memon, Mughal, Pathan, Qureshi, Rajput, Sheikh, Syed, and Yousafzai. All other castes showed a specific incidence of RGD. A detailed outlook of the RGD-caste relationship is shown in Figure 4.

### Association of Consanguinity with RGDs

There were four categories for recording the consanguinity status of the parents in RGD carrier families: first cousin, second cousin, relative, and unrelated. Among the RGD families, first cousin marriages were 129 (77.2%), second cousin marriages were 14 (8.4%), marriages within relatives were 7 (4.2%), and 17 (10.2%) couples were married to an unrelated spouse. Cumulatively, first cousins, second cousins, and marriages with relatives accounted for 89.8% of the current cohort in this registry. This is one of the highest consanguinity rates observed in RGD carrier families.

To illustrate the distribution of families affected by various RGDs according to consanguinity status, a stacked bar plot shows RGD-wise consanguinity rates in Figure 5. The visualization in Figure 5 highlights a pronounced trend in which a significant number of RGDs showcase consanguineous marriages, particularly first-cousin marriages. In our current registry cohort, 100% first-cousin marriages were observed in 50 RGDs, 100% second-cousin marriages were observed in four RGDs (methyl crotonyl-coA carboxylase deficiency, mitochondrial complete deficiency type 1, POLG-related disorder, and short rib thoracic dysplasia type 2), 100% marriage with a relative in one RGD (nephropathic cystinosis), and 100% marriage with an unrelated person in three RGDs (HHH syndrome, HADH-related disorder, and X-linked adrenoleukodystrophy). Thus, 69 out of the 72 RGDs in our registry showed a history of consanguinity, suggesting a causal role of consanguinity in the incidence of RGD in this cohort. A total of 14 RGDs showed mixed patterns of consanguinity. These RGDs include cystic fibrosis, citrullinemia, Gaucher disease, glutaric aciduria type 1, glycogen storage diseases, hemophilia, methylmalonic aciduria, MPS, NP, osteoporosis, PFIC, Pompe disease, Sandhoff disease, and spinal muscular atrophy. All these RGDs showed different percentages of consanguinity patterns, but the consanguinity rate remained 50% or higher, consistent with our observation of consanguinity in the causal role. To further assess the statistical association between consanguinity and RGDs, we used previously published consanguinity data of 1011 families from the Punjab Consanguinity Survey with no RGDs (Shenk et al., 2024) and compared them with our consanguinity records using a chi-square test (X^2^=44.78, p-value<0.00001). The association was extremely statistically significant, supporting our observation of the causal association of consanguinity with RGDs.

**Figure 5:**
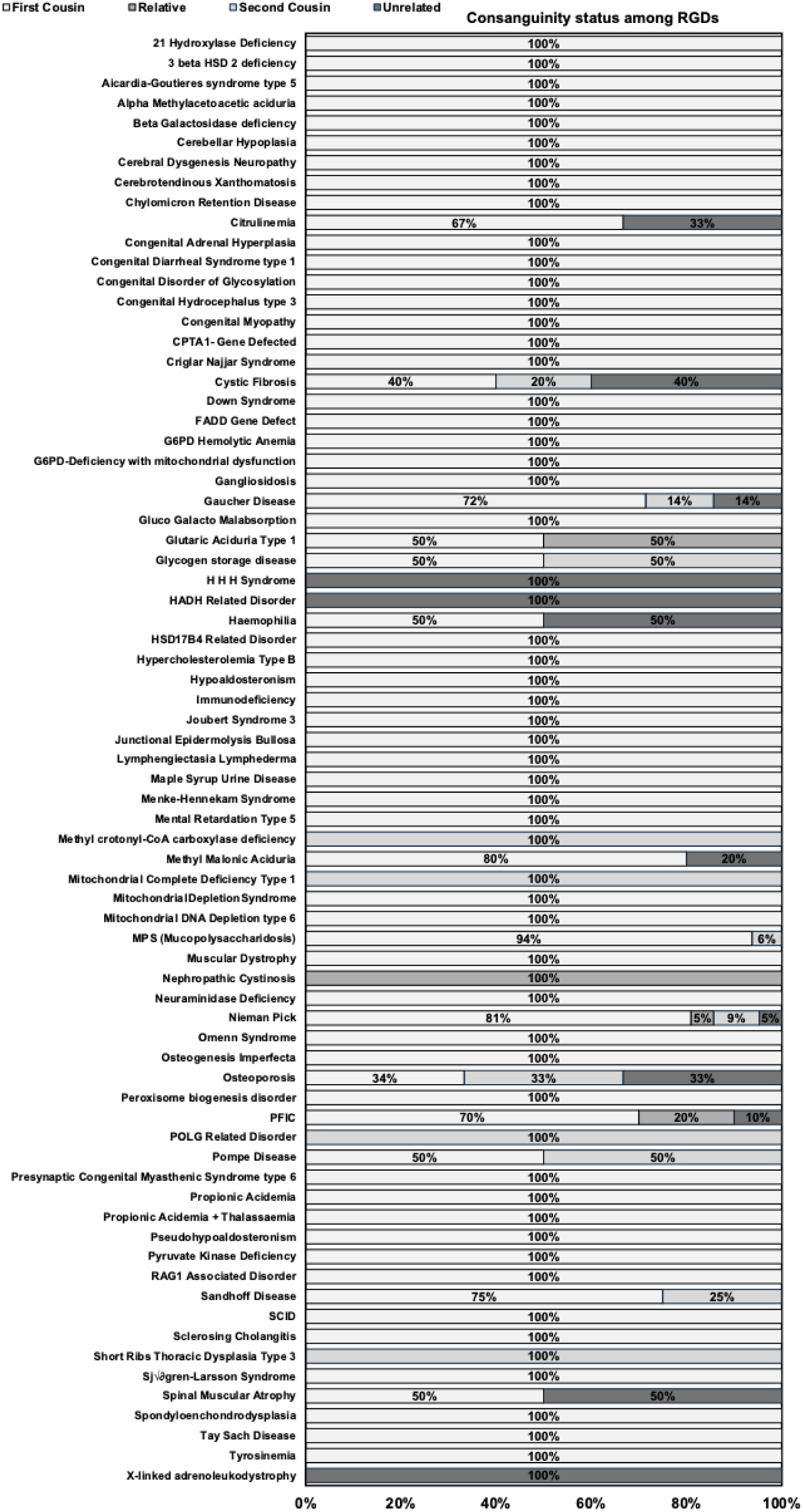
Consanguinity comparison among RGDs. Consanguinity status among RGD carrier families is presented. Consanguinity is recorded as first cousin marriage, second cousin marriage, marriage with a relative and marriage with unrelated spouse.

Significant variations in consanguinity patterns among different ethnic groups were also observed, and the cumulative share of each ethnic group is shown in Supplementary Figure 3. Punjabi, Pathan, Saraiki, and Sindhi shared both consanguinity and non-consanguinity patterns, while Kashmiri, Balochi, and Hindko showed only consanguinity patterns.

### Molecular epidemiology of RGDs in current registry

Following the registration and identification of RGDs in the current registry, we examined genetic variants identified by NGS in 91 RGD families spanning a repertoire of 110 genes, including primary and secondary RGD findings. Most RGDs and causal genes are unique to each family; however, there are RGDs in which the causal gene or pathogenic variant is observed in more than one family. The family-wise incidence of each RGD and gene repertoire is shown in Supplementary Table 2A. These 110 genes cumulatively had 131 variants, and disease-associated genes and variants with family incidence are listed in Supplementary Table 2B. The gene-wise relationship is presented in Supplementary Table 2C, while the variant-wise relationship is presented in Supplementary Table 2D.

For each of the 131 variants, we annotated rsIDs and allele frequencies from the dbSNP and gnomAD databases. These allele frequencies, along with rsIDs, are listed in Supplementary Table 2A. The allele frequencies in this table are categorized according to global ethnicities, including African, American, European, Middle Eastern, and South Asian ancestries. The purpose of this comparison was to determine whether the pathogenic variants in our dataset are population-specific or globally distributed. Although most pathogenic variants from our cohort were global, 24 pathogenic variants associated with 23 RGDs were specific to the South Asian population. This population specificity of pathogenic variants of associated RGDs argues if these variants are founded in the South Asian population, and termed as founding pathogenic variants. The allele frequencies of these variants, according to the gnomAD database, are shown in Figure 6A. These 23 RGDs with founding pathogenic variants include Aicardi-Goutières syndrome type 5, α-methyl acetoacetic aciduria, aspartyl glucosaminuria, breast cancer, Cerebrotendinous Xanthomatosis, congenital hypoaldosteronism, dihydro-pyrimidine-dehydrogenase deficiency, hemophagocytic lymphohistiocytosis, junctional epidermolysis bullosa, microencephaly, MPS I, MPS IV, neuraminidase deficiency, Omenn syndrome, ornithine transcarbamylase deficiency, PFIC II, Pompe disease, pontocerebellar hypoplasia type II, Sandhoff disease, severe combined immunodeficiency, spondyloenchondrodysplasia, Stromme syndrome and 3-β hydroxysteroid dehydrogenase type II deficiency. Among these diseases, the same MPS I founding variant in the IDUA gene was observed in three different families.

**Figure 6:**
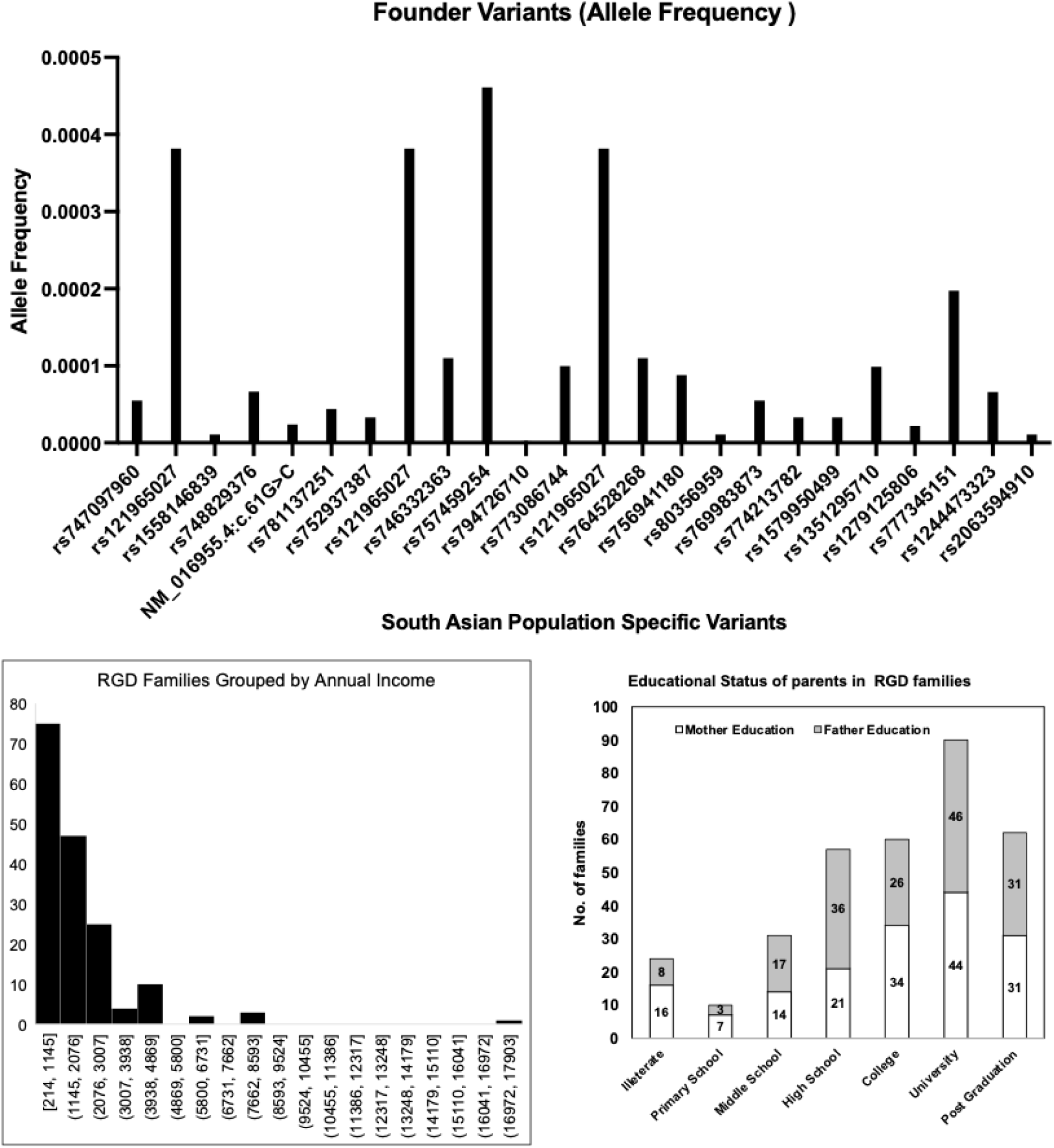
Highlights of demographics and founder variation in study cohort. A: Allele frequency of South Asian Founding Pathogenic Variants identified in current study B: Annual income of RGDs families in current study plotted in eight groups dividing low-, low-middle, high-middle and high income groups C: Educational status of RGD parents in current study exhibits major proportion of highly literate parents

In addition to the South Asian population specificity, we investigated variants with shared origins in the European and Middle Eastern populations. We found 19 variants with ancestry shared between South Asian and non-Finnish European populations only (Supplementary Table 2A). Moreover, we found two variants of shared ancestry between the South Asian and Middle Eastern populations according to the gnomAD allele frequency database. Four variants were shared among South Asian, European, and Middle Eastern populations. The higher shared variants observation between South Asian, Middle Eastern and European populations is consistent with Out-of-Africa migration history. It is also worth noting that both the current study population and the European population have Caucasian origins. Identification of founder pathogenic variants in this study is also suggestive of inclusion of underrepresented populations in global genomic and clinical projects.

### Association of Socioeconomic demographics with RGDs

The entire family of RGD patients is affected psychologically, socially, culturally, and economically. In this study, we conducted an in-depth evaluation of RGD carrier families to investigate whether they belong to the low-, middle-, or high-income groups. In the current cohort of our registry, the annual income of RGD carrier families ranged between $214 and $17142, with a mean annual income of $1780. An income group classification is shown in Figure 6B, where RGD families are divided into eight groups with an interval size of 931. We used the World Bank’s gross national income (GNI) per capita as the annual income per RGD carrier family to rank the economic status of each family. The first group, with an annual income between $214 and $1145 had 75 RGD carrier families, which is consistent with the World Bank’s low-income group, suggesting a high incidence of low-income families carrying RGDs. Second, there were 86 families with annual family incomes between $1145 and $4869, consistent with the World Bank’s low-middle income group. The third group, with an annual family income between $5800 and $8593, ranked as the high-middle income group, and had five RGD carrier families. The fourth group had only one family with an annual income of $17142 and ranked in the high-income group. In conclusion, the 161 families in the current cohort belonged to either the low-or lower-middle-income groups, implying a higher economic burden on RGD carrier families. While assuming a higher economic burden, it is necessary to remember that there is no national health insurance system in Pakistan; therefore, any costs associated with the patient are out-of-pocket expenses for the RGD family, creating inequities in accessing standard healthcare.

We further collected data on the literacy rate, including the highest education status of these 167 RGD families. Figure 6C shows the educational status among RGD families, with the highest number of spouses with at least university-level education, followed by post-graduation, college, and high school level education. This high literacy rate in this cohort is consistent with the growing literacy rate in Pakistan and provides a niche for genetic counselling and better care of RGD patients if appropriate resources are provided at the national or provincial scale.

## Discussion

This study is the first and only RGD registry with molecular, health, and socioeconomic data from Pakistan. In this manuscript, we report the spectrum of RGDs in the Punjabi population for the first time. Several individual case reports of RGDs in the Punjabi population have been published; however, a population-wide spectrum is not known. The PTGDPRI took the initiative to build a province-wide registry and registered 167 families with 72 RGDs of pediatric onset (Figure 1). These 72 RGDs include metabolic diseases, neurodevelopmental diseases, immunodeficiency diseases, connective tissue diseases, and others. In addition to disease identification, we annotated 110 genes with 106 RGD phenotypes, including primary and secondary diagnoses. These genes showed 91 primary variants and 58 secondary variants, of which 31 variants among these were VUS. Furthermore, we identified RGDs among Punjabi and non-Punjabi ethnic groups, and a caste-wide incidence was also presented. In our registry, we also incorporated consanguinity as a risk factor for RGDs and found that 89.8% of RGD carrier families were consanguineous, and an association between RGD and consanguinity was observed. Molecular epidemiology methods suggested that 24 of the total registered variants were specific to South Asian populations and were annotated as the founding pathogenic variants. Finally, we observed an economic burden giving rise to inequities in accessing healthcare despite a high literacy rate among registered RGD families.

As mentioned earlier, PTGDPRI, a provincial government institute known for genetic testing of thalassemia and other genetic diseases, came across the line of action to prevent such RGDs that are life-threatening and affect a large population of the Punjab region. In the dataset presented in this manuscript, 167 RGD carrier families were registered in PTGDPRI and obtained clinical services, including CVS sampling, prenatal genetic testing, and genetic counselling, based on their clinical picture or past clinical history; thus, the majority of registered families were already identified as risk groups. In the current dataset, registered families had 293 affected children, including 188 who had already died, and only 98 children were born normal. We have calculated burden of RGD-affected children in these families and mean RGD-burden was 0.81±0.24 out of 1.00 presenting a significant RGD-affected children burden on each family (Figure 2A). Although several previous studies have reported the incidence of genetic diseases in the Pakistani and Punjabi populations, this evidence remains generalized to phenotypes, traits, and common genetic diseases such as hemoglobinopathies (Bibi et al., 2022, Naqvi et al., 2024, Azmatullah et al., 2024, Bhatti et al., 2019, Nawaz et al., 2025, Ahmad and Naeem, 2025). In addition to these traits and common genetic diseases, there have been many case reports of RGDs, but collective evidence of a population-wide spectrum is lacking. This is the first report of clinically diagnosed RGDs based on molecular evidence. RGD patients are mostly children, with associated higher morbidity and mortality, and the worst of these is the diagnostic odyssey (Mazzucato et al., 2023, Ferreira, 2019). The RGD carrier families in our registry represent the final outcome of any diagnostic odyssey they faced in the past. This registry not only compiles epidemiological information on RGDs but also serves as a platform to introduce carrier families with their desired clinical services through its field presence in the Province of Punjab.

In our registry, we found that metabolic RGDs were the most prevalent, followed by neurodevelopmental, immunodeficiency, congenital, endocrine, connective tissue, lymphatic, skeletal, bone, and hematological RGDs. This is the first population-based evidence to understand the RGD trends. Among the high-incidence metabolic RGDs, 37 were inborn errors of metabolism. Although these metabolic RGDs, including inborn errors of metabolism, are fatal, many of them are curable after the identification of metabolic malfunction or metabolic element deficiency (Gambello and Li, 2018). This potential cure highlights the need for fast and accurate genetic testing availability to the general population and/or at least to carrier families which is currently a challenge for RGD carrier families.

In this study, the five most common RGDs were NP, PFIC, MPS, Gaucher disease, and methylmalonic aciduria. Together, these diseases account for approximately 40% of the RGD burden in our registry. In addition to the higher incidence of RGDs, we also observed varying phenotypes of NP, PFIC, and MPS. These phenotypes included NP I and NPC for NP; PFIC I, PFIC II, PFIC IV, and PFIC V for PFIC; and MPS I, MPS IIIA, MPS IIIB, and MPS IV for MPS. In addition to these RGDs, many other families exhibit several secondary variants in addition to the primary variants. The existence of secondary variants is evidence of the unique genotype of each RGD patient, which then produces a unique phenotype of the clinical condition. In addition, these molecular findings of unique genotypes remain consistent for associated primary and secondary genes and pathways; thus, a pathway-based approach can also be established for the identification of RGDs at the molecular level. Current diagnosis best practices mainly include WGS, WES or targeted sequencing which are not always successful due to complexities associated with RGDs and technical limitations of these technologies. However, involvement of multiple genes suggests disruption of physiological pathways can be used in a reverse fashion to identify associated genes or at least understand molecular scale pathology of RGDs. The PTGDRI currently only offers prenatal testing and genetic counselling, but a complete solution with intervention is desired by provincial or national stakeholders. Several treatment regimens with regulatory approval are already available for these RGDs, and immediate relief in terms of accessible and equitable clinical services is necessary (Santos-Lozano et al., 2015, Matencio et al., 2020, Loomes et al., 2022, Gunaydin and Bozkurter Cil, 2018, McKiernan et al., 2024, Kingma and Jonckheere, 2021, Stapleton et al., 2019). In addition to the available treatment regimens, many potential therapeutics are being developed and are under clinical trials. Access to potential therapeutics for these patients is also necessary for clinical trial enrollment.

Furthermore, it is not only about RGDs; we have tried to incorporate the maximum available information associated with RGDs. The impact of carrying RGD-associated alleles does not only affect newborn children, but also worsens the family well-being and reproductive health of carrier mothers, as shown by higher child mortality and GPA statistics in our registry. We learned that the families in our registry had 2.34±1.1 total children, with only 0.59±0.7 normal children. The remaining children were either affected or deceased. The mean mortality burden per family in our registry was 1.13±1.1, which is approximately half of the total mean children in each family. These data suggest a child mortality rate of 480 per 1000, which is significantly higher than the national average of 62 children per 1000, arguing that higher mortality rates (seven times the national average) are associated with RGDs (Demographic, Shenk et al., 2024). These higher numbers of mortality and RGD-affected children worsened family well-being, although we did not investigate this on an established scale. In addition to higher mortality, RGD carrier mothers also face a higher incidence of abortion. The GPA statistics (Figure 2A) indicated that the mean G was 3.92, with mean P, 2.34 and mean A, 0.58. This P is also representative of the fertility rate among RGD carrier mothers, which is much lower than the national fertility rate of 3.6 per mother (Demographic, Shenk et al., 2024), arguing lower fertility rates than the national average. The difference between P and the other two statistics, G and A, represents the fertility challenges faced by RGD carrier mothers and families, especially in terms of lower viable births and higher rate of abortions. Although this trend of abortions was not common among all RGDs, 37 RGDs showed an incidence of A in 66 RGD families (Figure 2B). In Pakistan, several maternal health surveys have revealed population rates of A between 29 and 66 per 1000 women (Khalid et al., 2025, Sathar et al., 2025, Sathar et al., 2007). However, we believe that our current dataset is still too small to draw conclusions about the abortion rate in RGDs in the study population. However, based on our results, it is evident that at least 37 RGDs are associated with abortion, and 66 carrier mothers out of 167 (395 per 1000 women, at least six times the national rate) had experienced an abortion at least once (Figure 2).

The Punjabi population and other ethnicities living in Pakistan are anthropologically and genetically heterogeneous (Malik and Amin-ud-Din, 2013, Aftab et al., 2017, Sajid Hussain et al., 2013), representing historical migrations to this region (de Gila-Kochanowski, 1990, Agarwal, 2000, Witzel, 2019, Pathak et al., 2018). Therefore, we profiled RGDs according to the major ethnic groups represented in our registry. These groups include Punjabi, Pathan, Saraiki, Sindhi, Kashmiri, Balochi, and Hindko families. Although all families were recruited from Pakistan, these ethnic groups are also widely distributed across other South Asian countries. The Punjabi population resides primarily in the Punjab province of eastern Pakistan and in the adjoining Indian state of Punjab. The Pathan (Pashtun) population mainly inhabits the northwestern province of Khyber Pakhtunkhwa in Pakistan and the neighboring regions of Afghanistan. The Saraiki population is concentrated in southern Punjab, Pakistan, and extends into the Indian Punjab. The Sindhi population lives in Pakistan’s Sindh province and the adjacent regions of Rajasthan and Gujrat in India. The Kashmiri people inhabit the Kashmir region, which is divided between India and Pakistan, while the Balochi population originates from Pakistan’s western province of Balochistan and extends into Iran and parts of Western and Central Asia. Thus, knowledge in this registry about ethnic groups is relevant not only to the Pakistani population but also to neighboring populations in South, West, and Central Asia. In the current registry version, we have 118 families of Punjabi origin, 34 Pathan, five Saraiki, four Sindhi, three Kashmiri, two Balochi, and one Hindko. Among these registered families, 58 RGDs were observed, of which 47 were Punjabi population-specific. Among the registered Pathan families, 16 RGDs were present, of which eight were Pathan population-specific. Among the Saraiki-registered families, five RGDs were present, two of which were Saraiki population-specific. There were only two Balochi families in this registry exhibiting two RGDs, one of which was Balochi population-specific. However, the Hindko and Kashmiri populations shared RGDs with the Punjabi and Pathan populations, and no population-specific RGDs were identified in this registry. Although these findings highlight the population specificity of several RGDs, there is still a chance that these RGDs could occur in other ethnic population groups. Because these registry data are more observational and do not lead to the inference that other populations could not be affected by these traits unless a genomic survey of other populations for pathogenic variants is conducted.

To narrow down this ethnic analysis, we stratified the data in our registry by caste. Castes are anthropologically isonym groups that tell us the lineage of each individual and are used for identification purposes even in modern-day South Asian societies. In some non-Punjabi ethnicities, this lineage information is preserved with tribe names instead of castes. However, in this registry, we used both terminologies, caste and tribe, synonymously. Among castes, more than 50% of the RGD burden was found in five Punjabi castes (Arain (13.2%), Rajput (13.2%), Sheikh (8.4%), Butt (5.4%), and Jutt (5.4%)) and one non-Punjabi group (Pathan (5.4%)). Other castes with lower RGD incidence include Baloch, Bhatti, Malik and Syed. The castes with higher RGD incidence, including Arain, Rajput, and Jutt, are large caste groups widely present in the Pakistani and Indian Punjab regions, Sindh province of Pakistan, and Haryana region of India. Moreover, these castes also include tens of subcastes that are representative of each group’s anthropology, region, trade, or religion, while keeping the caste-wide history intact. Other castes, including Sheikh, Butt, and Pathan, are also widely present in Pakistan, India, present-day Kashmir, and Afghanistan, but their populations are smaller than the aforementioned castes. In our previous studies and other published literature, we learned that the incidence of several genetic traits was higher in Arain, Rajput, Jutt, and Sheikh castes (Akhtar et al., 2019, Bibi et al., 2022, Naqvi et al., 2024, Azmatullah et al., 2024, Bhatti et al., 2019, Nawaz et al., 2025, Ahmad and Naeem, 2025, Aslamkhan et al., 2023). Thus, our current registry is a valuable resource for caste-specific RGDs and could be useful for RGD screening of specific castes. In the current registry, we found that 51 RGDs were caste-specific. Rajput showed seven caste-specific RGDs, Arain six, Sheikh five, Jutt three, and Butt two caste-specific RGDs. Our results on ethnicity and caste-specificity suggest the need to build a comprehensive registry of RGDs among South Asian ethnicities and castes. Such a registry could provide not only comprehensive information on ethnic and caste-specificity but also generate screening batteries for groups of RGDs. This is also a goal of the current registry to build a comprehensive resource and screening battery for a group of RGDs, which could be a quick and cost-efficient way to screen RGD-suspects before genetic testing and/or genomic profiling, especially in South Asian countries, where access to modern genetic technologies is still limited.

Following castes, we led our registry to another aspect of the social structure, consanguinity. In a society divided by castes, people prefer to interbreed within their caste as it is a matter of pride for their families. In addition, consanguinity is a practice to maintain social, political, and economic status within the family. Though a few religions in South Asia including Sikhism in Punjabi population prohibit consanguineous marriages but people practicing Islam vastly practice it not only in South Asian premises but also in Middle Eastern and Western and Central Asian regions (Akhtar et al., 2019, Zar et al., 2020, Ghanim et al., 2023, Mofied et al., 2024, El Goundali et al., 2022, Alshaban et al., 2025, Popescu et al., 2025, Mansouritorghabeh, 2021). In our registry, 69 out of 72 RGDs showed history of consanguinity practice (Figure 5). In our previous population-based surveys, we found incidence of consanguinity is over 80% in Punjab (Akhtar et al., 2019). In current registry, we observed 89.8% in RGD carrier families with 77.2% first cousin marriages. This is so far the highest consanguinity burden reported in any population in the region. As several previous studies has tested consanguinity as a risk factor for recessive genetic traits, so we have tested consanguinity status in our registry against non-RGD data of Punjabi population previously available from Shenk et al (Shenk et al., 2024). This analysis revealed a significant association between RGDs and consanguinity and consanguinity remains the major contributor of RGDs in current RGD carrier families. Identification of RGD-and other lethal genetic trait-carriers pre-marital can serve as control strategy of RGDs inheritance to next generation in this high consanguinity practice. The major contributors to consanguinity as mentioned earlier are social, economic and political reasons. So, we have investigated socioeconomic demographic factors including annual income and education status among RGD carrier families in this registry. We found 75 RGD carrier families to be among low-income group, 86 among low-middle income groups, five among high-middle income and only one high income family (Figure 6). This suggests a huge economic pressure and lack of financial resources for diagnosis and treatment of RGDs. Such economic pressures and long-standing diagnostic odyssey previously have been reported in Pakistani population, several developed nations and under-privileged populations (Akesson et al., 2025, Glaubitz et al., 2025, Zaman et al., 2025). This is also worth noting that unlike global north countries, no national health insurance or coverage system exists in Pakistan and all clinical costs are out-of-pocket ultimately an economic burden for these families despite existing economic pressure. These data suggest that in our current study population, the share of socioeconomic status and wealth may not be the primary reason for consanguinity but rather the lack of options for marriage partnerships due to economic stress. The Punjab Consanguinity Survey (Shenk et al., 2024) also revealed similar outcomes, where the economic burden on families is among the major contributor of consanguinity practice in the Punjabi population. Despite the high economic pressure on RGD families, we found that the literacy rate among these RGD families was very high, which also reflects the socioeconomic dilemma of low wages and unemployment in Pakistan and other developing countries. However, in the current study, highly literate people are good resources for understanding RGDs and seeking genetic counselling.

Finally, we investigated the global allele frequencies of pathogenic variants in the current registry. The allele frequencies of pathogenic variants suggest the prevalence and incidence of particular genotypes and associated disease phenotypes. In our registry, we identified 131 variants in 110 genes associated with RGDs. We searched for their incidence among all major population groups and learned that 24 of these variants were of South Asian origin and were termed founding pathogenic variants. These 24 variants were associated with 23 RGDs, and these variants should be prioritized and incorporated into genetic tests and pan-genomic testing panels for RGDs for a fast, cheaper, and accurate diagnosis, minimizing the present odyssey. In addition, structural changes induced by these founder pathogenic variants should be addressed in ongoing novel drug discovery efforts and companion diagnostics.

In addition to the direct results of our study, there are several indirect points of discussion, including the need for rapid and cost-efficient genetic testing and the need for provincial, national, and regional RGD support groups. The current registry is an effort by the PTGDPRI to understand RGDs dynamics, but the following efforts are needed by the provincial government of Punjab, the national ministry of health coordination, and the World Health Organization’s regional office of the Eastern Mediterranean Region and Southeast Pacific region. Pakistan is representative of a diverse, heterogeneous population with cultural roots shared between Southeast Asia and the Eastern Mediterranean Region. Several aspects reported in this study, including lower fertility rates among RGD carrier mothers, higher mortality rates among RGD carrier families, higher incidence of consanguinity in RGD carrier families, and identification of founding variants, lay the foundation for policy-making in both regions. To better manage RGDs, national-level infrastructure, including newborn and prenatal screening of RGDs, is necessary and should be mandated as soon as possible by key players, including non-government and not-for-profit organizations.

Another aspect that is primarily missing in our results, but is a translational aspect of our results, is the need for rapid and cheap diagnostic assays for RGDs. The current diagnosis gold standard for RGDs is either WGS or WES, but their cost is not bearable for patients in the South Asian region and some regions of the Eastern Mediterranean Region. Thus, based on the shared ancestry and genetic admixture in these regions, we suggest building a regional database of genetic variants. This database could be useful for cheaper and rapid RGD diagnosis in the region. As the region-specific information is much clearer by our discovery of tens of founding pathogenic variants, other previous studies (Tournebize et al., 2022, Kerdoncuff et al., 2025) have also reported it. Therefore, we suggest building a broader regional catalog of pathogenic and founder pathogenic variants, and these efforts should be accelerated by financial and scientific incubations. Moreover, a subsidy should be announced for organizations, companies, and startups that can take up the challenge of building cheaper diagnostics. These could be genotype arrays, multiplexed PCRs, cheaper and rapid sequencing methods, population-specific or pan-genomic gene panels. In addition, following diagnosis, genetic counselling and efforts to ensure treatment availability are needed. All these things together could be compiled in one RGD policy, which is immediately needed in Punjab and Pakistan and also in the whole region.

This registry provides the first foundational dataset for RGDs in the Punjabi population and represents a model for integrating molecular epidemiology with public health genetics. The Punjabi population of Pakistan illustrates how genetic, socioeconomic, and cultural factors converge to amplify the burden of RGDs in this population. High consanguinity, limited diagnostic access, and the absence of policy infrastructure act synergistically to sustain this burden in Pakistan. There is an urgent need to develop cost-effective diagnostic assays that target RGD-associated variants prevalent in South Asian and Middle Eastern populations to minimize inequity to access healthcare. Translating advanced molecular technologies into affordable diagnostic platforms is crucial for achieving this goal. Expansion of this registry into a national genomic surveillance program could enable early detection, equitable data sharing, and transnational research collaborations. Comparative allele frequency analyses using the gnomAD database further demonstrated that several pathogenic variants are exclusive to South Asian ancestry, underscoring regional founder effects and the critical need to include underrepresented populations in global RGD genomics.

## Supporting information

List of supplementary tables

## Data Availability

All data produced in the present work are contained in the manuscript.

## Acknowledgement

We gratefully acknowledge the PTGDPRI for their collaboration and provision of clinical and genetic data used in this study. We extend our sincere thanks to the families and patients who generously consented to participate in this study.

This study is based on the work conducted as part of the doctoral dissertation of the first author at the CEMB, PU. We also thank the technical and administrative staff at PTGDPRI for their assistance in patient coordination and data management and CEMB, PU, for providing computational and analytical resources that supported data analysis.

This study did not receive any dedicated external funding but was conducted using institutional resources from the University of California, Davis, and The University of Tokyo.

## Author contributions

Conceptualization, I.T. and M.S.A.; Methodology, I.T. and M.S.A; Software, M.S.A; Validation, M.S.A and M.S.; Formal Analysis, I.T.; Investigation, I.T.; Resources, M.S.A.; Data Curation, I.T.; Writing – Original Draft Preparation, I.T. and M.S.A.; Writing – Review & Editing, I.T., M.S.A. and M.S.; Visualization, I.T.; Supervision, M.S.A and M.S.; Project Administration, M.S.

---

**Supplementary Figure 1:**
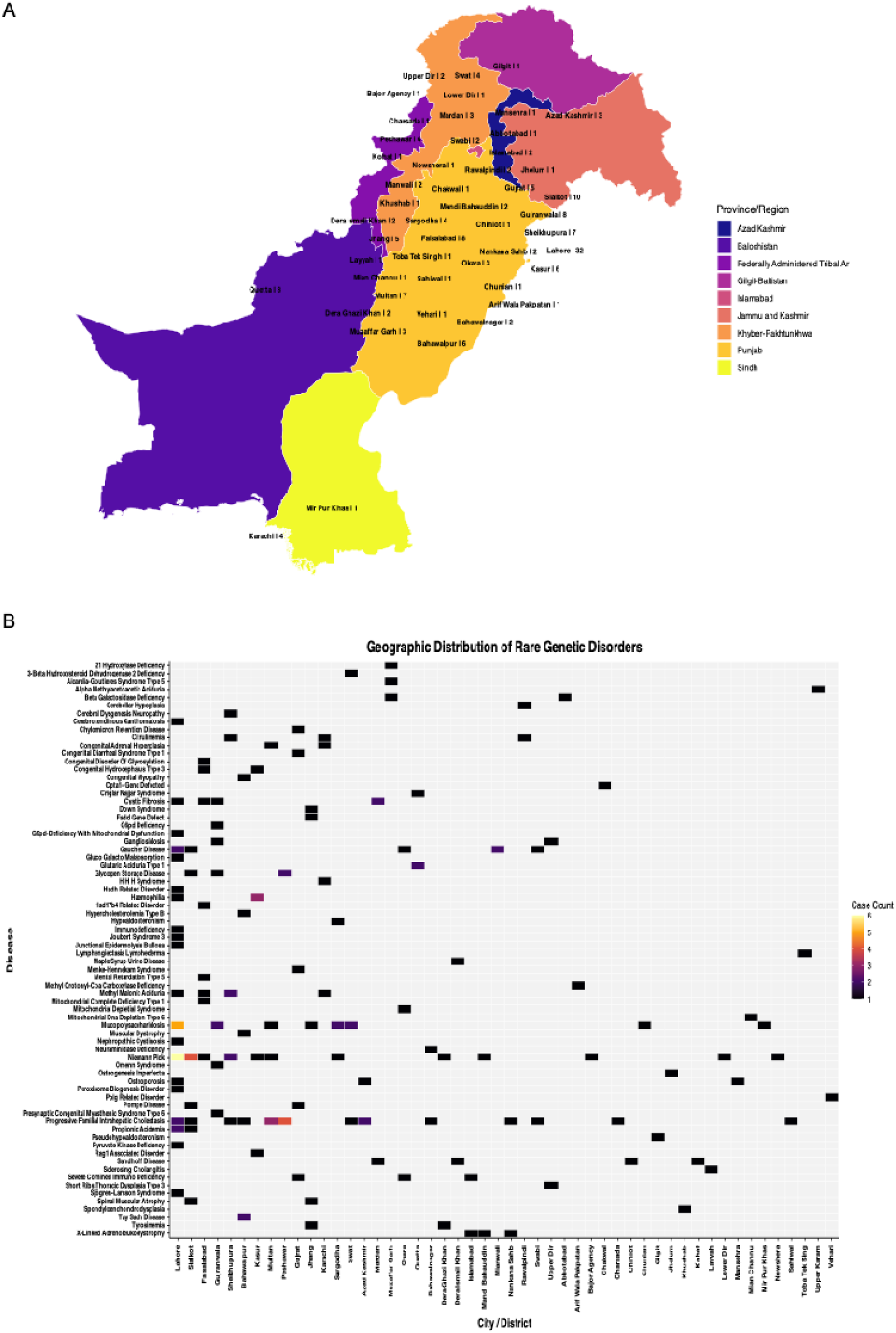
Geographical distribution of RGDs. A: The mapping of RGD families across the Punjab Province illustrates the spatial distribution of families affected by RGDs, highlighting regional clustering and population-specific concentration patterns B: The geographical distribution of RGDs in various districts showing areas with higher and lower disease occurrence

**Supplementary Figure 2:**
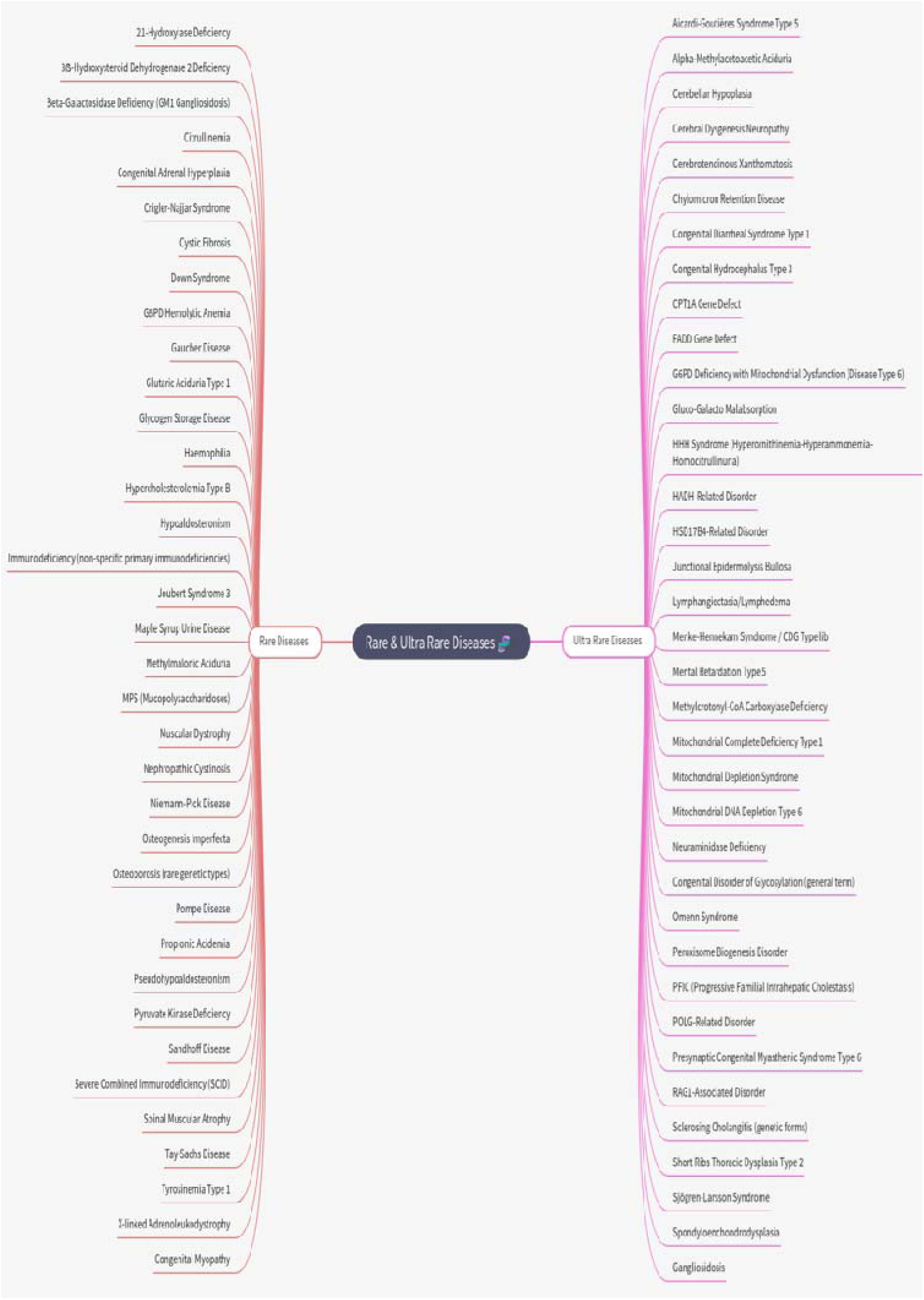
Classification of rare and ultra-rare diseases. This figure illustrates classification of rare & ultra-rare diseases according to their occurrence rates, distinguishing rare diseases from ultra-rare diseases to better understand their epidemiological and clinical distribution

**Supplementary Figure 3:**
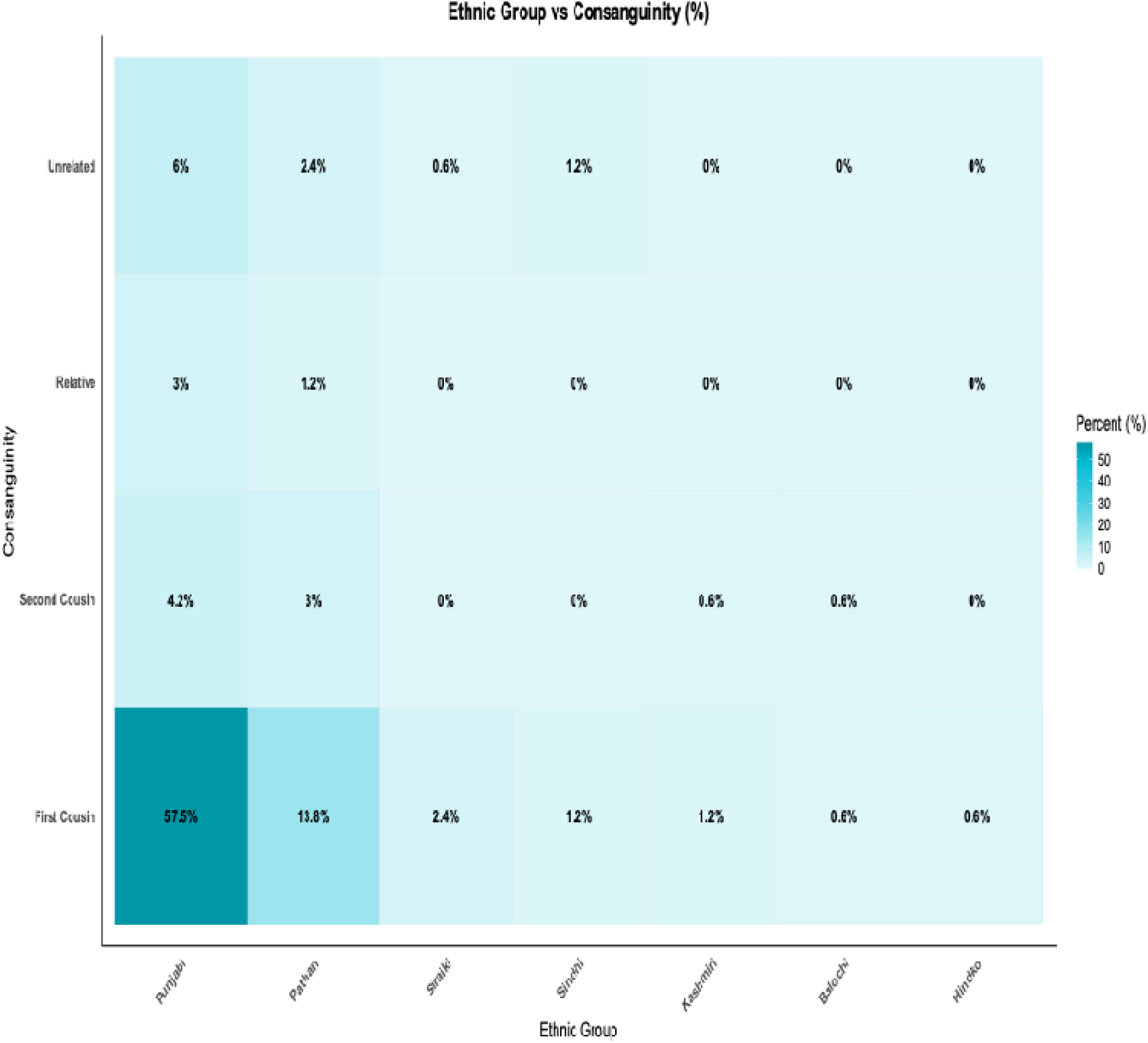
Consanguinity comparison among Ethnic Groups. This pattern shows a comparison of consanguinity and ethnicity among families affected by RGDs showing variation in consanguineous marriage rates across ethnic groups

## References

100 , G. P. P. I. 2021. 100,000 genomes pilot on rare-disease diagnosis in health care—preliminary report. New England Journal of Medicine, 385, 1868–1880.

2. Abudejaja, A., Khan, M. A., Singh, R., Toweir, A. A., Narayanappa, M., Gupta, B. & Umer, S. 1987. Experience of a family clinic at Benghazi, Libya, and sociomedical aspects of its catchment population. Family practice, 4, 19–26.

3. Acharya, S. & Sahoo, H. 2021. Consanguineous Marriages in India: Prevalence and Determinants. Journal of Health Management, 23, 631–648.

4. Adeyemo, A. A., Amodu, O. K., Ekure, E. E. & Omotade, O. O. 2018. Medical genetics and genomic medicine in Nigeria. Molecular genetics & genomic medicine, 6, 314.

5. Adhikari, A. N., Gallagher, R. C., Wang, Y., Currier, R. J., Amatuni, G., Bassaganyas, L., Chen, F., Kundu, K., Kvale, M. & Mooney, S. D. 2020. The role of exome sequencing in newborn screening for inborn errors of metabolism. Nature medicine, 26, 1392–1397.

6. Aftab, H., Ambreen, A., Jamil, M., Garred, P., Petersen, J. H., Nielsen, S. D., Bygbjerg, I. C. & Christensen, D. L. 2017. High prevalence of diabetes and anthropometric heterogeneity among tuberculosis patients in Pakistan. Tropical Medicine & International Health, 22, 465–473.

7. Agarwal, V. 2000. What is the Aryan Migration Theory?

8. Ahluwalia, J. K., Hariharan, M., Bargaje, R., Pillai, B. & Brahmachari, V. 2009. Incomplete penetrance and variable expressivity: is there a microRNA connection? Bioessays, 31, 981–992.

9. Ahmad, R. & Naeem, M. 2025. A systematic review of hereditary neurological disorders diagnosed by whole exome sequencing in Pakistani population: updates from 2014 to November 2024. Neurogenetics, 26, 40.

10. Akesson, L. S., Parekh, S., Alderdice, A., Jackson, H., Bain, L., Dudgeon, A., Williamson, L. J., Akesson, B. L., Say, G., Kellett, M. J., POPE-Couston, R. & Wallis, M. J. 2025. Developing best practice rare disease diagnostic care models in a real-world rural/regional setting. medRxiv, 2025.07.24.25332141.

11. Akhtar, M. S., Ashino, R., Oota, H., Ishida, H., Niimura, Y., Touhara, K., Melin, A. D. & Kawamura, S. 2022. Genetic variation of olfactory receptor gene family in a Japanese population. Anthropological Science, 211024.

12. Akhtar, M. S., Aslamkhan, M., Zar, M. S., Hanif, A. & Haris, A. R. 2019. Dichromacy: Color Vision Impairment and Consanguinity in Heterogenous Population of Pakistan. The International Journal of Frontier Sciences, 3, 41–56.

13. Akthar, M. S. 2019. Role of epidemiological studies in disease prevention. The International Journal of Frontier Sciences, 3, 1–2.

14. Alqahtani, A. S., Alotibi, R. S., Aloraini, T., Almsned, F., Alassali, Y., Alfares, A., Alhaddad, B. & AL Eissa, M. M. 2023. Prospect of genetic disorders in Saudi Arabia. Frontiers in Genetics, 14, 1243518.

15. Alshaban, F. A., Aldosari, M., Ghazal, I., AL-Shammari, H., Elhag, S., Thompson, I. R., Bruder, J., Shaath, H., AL-Faraj, F., Tolefat, M., Nasir, A. & Fombonne, E. 2025. Consanguinity as a Risk Factor for Autism. Journal of Autism and Developmental Disorders, 55, 1945–1952.

16. Aslamkhan, M., Ali, A. & Barnett, H. 1969. Consanguineous marriages in rural west Pakistan. Ann Rept Uni Md Sch Med Icmrt, 69, 181–92.

17. Aslamkhan, M., Qadeer, M. I., Akhtar, M. S., Chudhary, S. A., Mariam, M., Ali, Z., Khalid, A., Irfan, M. & Khan, Y. 2023. Cultural consanguinity as cause of β-thalassemia prevalence in population. medRxiv, 2023.06. 01.23290856.

18. Auerbach, B. J., Hu, J., Reilly, M. P. & Li, M. 2021. Applications of single-cell genomics and computational strategies to study common disease and population-level variation. Genome Research, 31, 1728–1741.

19. Aymé, S. 2012. State of the art of rare disease activities in Europe: a EUCERD perspective. Orphanet Journal of Rare Diseases, 7, A1.

20. Azmatullah, Khan, M. Q., Jan, A., Mehmood, J. & Malik, S. 2024. Prevalence-pattern of congenital and hereditary anomalies in Balochistan Province of Pakistan. Pak J Med Sci, 40, 1898–1906.

21. Baldovino, S., Sciascia, S., Carta, C., Salvatore, M., Cellai, L. L., Ferrari, G., Lumaka, A., Groft, S., Alanay, Y. & Azam, M. 2025. A global survey about undiagnosed rare diseases: perspectives, challenges, and solutions. Frontiers in Public Health, 13, 1510818.

22. Bazalar-Montoya, J., Cornejo-Olivas, M., Duenas-Roque, M. M., Purizaca-Rosillo, N., Rodriguez, R. S., Milla-Neyra, K., De la torre-Hernandez, C. A., Sarapura-Castro, E., Galarreta Aima, C. I. & Manassero-Morales, G. 2024. Clinical genome sequencing in patients with suspected rare genetic disease in Peru. NPJ Genomic Medicine, 9, 51.

23. Bener, A. & Alali, K. A. 2006. Consanguineous marriage in a newly developed country: the Qatari population. Journal of biosocial science, 38, 239–246.

24. Bhatti, N. A., Mumtaz, S. & Malik, S. 2019. Epidemiological study of congenital and hereditary anomalies in Sialkot District of Pakistan revealed a high incidence of limb and neurological disorders. population, 7, 9.

25. Bibi, A., Naqvi, S. F., Syed, A., Zainab, S., Sohail, K. & Malik, S. 2022. Burden of Congenital and Hereditary Anomalies in Hazara Population of Khyber Pakhtunkhwa, Pakistan. Pak J Med Sci, 38, 1278–1284.

26. Birkelund, C. H. 2019. Rare disease thresholds-An analysis of different definitions, laws and arguments.

27. Bittles, A. H. & Black, M. L. 2010. Consanguinity, human evolution, and complex diseases. Proceedings of the National Academy of Sciences, 107, 1779–1786.

28. Bizzari, S., Nair, P., Hana, S., Deepthi, A., AL-Ali, M. T., AL-Gazali, L. & EL-Hayek, S. 2023. Spectrum of genetic disorders and gene variants in the United Arab Emirates national population: insights from the CTGA database. Frontiers in Genetics, 14, 1177204.

29. Black, N., Martineau, F. & Manacorda, T. 2015. Diagnostic odyssey for rare diseases: exploration of potential indicators. Policy Innovation Research Unit (PIRU).

30. Boyarchuk, O., Kostyuchenko, L., Akopyan, H., Bondarenko, A., Volokha, A., Hilfanova, A., Savchak, I., Nazarenko, L., Yarema, N. & Urbas, O. 2024. Nijmegen breakage syndrome: 25-year experience of diagnosis and treatment in Ukraine. Frontiers in Immunology, 15, 1428724.

31. Carmichael, N., Tsipis, J., Windmueller, G., Mandel, L. & Estrella, E. 2015. “Is it going to hurt?”: the impact of the diagnostic odyssey on children and their families. Journal of Genetic Counseling, 24, 325–335.

32. Cerkauskaite-Kerpauskiene, A., Navickaite, M., Savige, J., Mazur, G., Brazdziunaite, D., Azukaitis, K., Slazaite, G., Laurinavicius, A., Miglinas, M. & Vainutiene, V. 2025. Lithuanian Study on COL4A3 and COL4A4 Genetic Variants in Alport Syndrome: Clinical Characterization of 52 Individuals from 38 Families. International Journal of Molecular Sciences, 26, 7639.

33. Charoute, H., Nahili, H., Abidi, O., Gabi, K., Rouba, H., Fakiri, M. & Barakat, A. 2014. The Moroccan Genetic Disease Database (MGDD): a database for DNA variations related to inherited disorders and disease susceptibility. European Journal of Human Genetics, 22, 322–326.

34. Chekroun, I., Rabea, F., Jain, R., Alsheikh-Ali, A. & Abou Tayoun, A. 2025. Premarital genomic screening in Arab populations of the Middle East. Nature Medicine, 31, 364–365.

35. Chung, C. C. Y., Project, H. K. G., Chu, A. T. W. & Chung, B. H. Y. 2022. Rare disease emerging as a global public health priority. Frontiers in public health, 10, 1028545.

36. Cipriani, V., Vestito, L., Magavern, E. F., Jacobsen, J. O. B., Arno, G., Behr, E. R., Benson, K. A., Bertoli, M., Bockenhauer, D., Bowl, M. R., Burley, K., Chan, L. F., Chinnery, P., Conlon, P. J., Costa, M. A., Davidson, A. E., Dawson, S. J., Elhassan, E. A. E., Flanagan, S. E., Futema, M., Gale, D. P., García-Ruiz, S., Corcia, C. G., Griffin, H. R., Hambleton, S., Hicks, A. R., Houlden, H., Houlston, R. S., Howles, S. A., Kleta, R., Lekkerkerker, I., Lin, S., Liskova, P., Mitchison, H. H., Morsy, H., Mumford, A. D., Newman, W. G., Neatu, R., O’toole, E. A., Ong, A. C. M., Pagnamenta, A. T., Rahman, S., Rajan, N., Robinson, P. N., Ryten, M., Sadeghi-Alavijeh, O., Sayer, J. A., Shovlin, C. L., Taylor, J. C., Teltsh, O., Tomlinson, I., Tucci, A., Turnbull, C., VAN Eerde, A. M., Ware, J. S., Watts, L. M., Webster, A. R., Westbury, S. K., Zheng, S. L., Caulfield, M. & Smedley, D. 2025. Rare disease gene association discovery in the 100,000 GenomesProject. Nature.

37. Clark, M. M., Hildreth, A., Batalov, S., Ding, Y., Chowdhury, S., Watkins, K., Ellsworth, K., Camp, B., Kint, C. I. & Yacoubian, C. 2019. Diagnosis of genetic diseases in seriously ill children by rapid whole-genome sequencing and automated phenotyping and interpretation. Science translational medicine, 11, eaat6177.

38. Cooper, D. N., Krawczak, M., Polychronakos, C., Tyler-Smith, C. & Kehrer-Sawatzki, H. 2013. Where genotype is not predictive of phenotype: towards an understanding of the molecular basis of reduced penetrance in human inherited disease. Human genetics, 132, 1077–1130.

39. Czech, M., Baran-Kooiker, A., Atikeler, K., Demirtshyan, M., Gaitova, K., Holownia-Voloskova, M., Turcu-Stiolica, A., Kooiker, C., Piniazhko, O. & Konstandyan, N. 2020. A review of rare disease policies and orphan drug reimbursement systems in 12 Eurasian countries. Frontiers in public health, 7, 416.

40. De Gila-Kochanowski, V. 1990. Aryan and Indo-Aryan Migrations. Diogenes, 38, 122–145. Demographic, P. Health Survey Demographic 2017–18.

41. Dharssi, S., Wong-Rieger, D., Harold, M. & Terry, S. 2017. Review of 11 national policies for rare diseases in the context of key patient needs. Orphanet journal of rare diseases, 12, 1–13.

42. Domaradzki, J. & Walkowiak, D. 2023. Ultra-rare ultra-care: The unique burden of ultra rare disease caregiving. European Journal of Paediatric Neurology, 48, 78–84.

43. Drugan, C., Procopciuc, L., Jebeleanu, G., Grigorescu-Sido, P., Dussau, J., Poenaru, L. & Caillaud, C. 2002. Gaucher disease in Romanian patients: incidence of the most common mutations and phenotypic manifestations. European Journal of Human Genetics, 10, 511–515.

44. Dumic, K. K., Grubic, Z., Kusec, V., Braovac, D., Gotovac, K., Vinkovic, M., Vucinic, M. & Dumic, M. 2023. The prevalence and genotype of 21-hydroxylase deficiency in the Croatian Romani population. Frontiers in Endocrinology, 14, 1170449.

45. EL Goundali, K., Chebabe, M., ZAHRA Laamiri, F. & Hilali, A. 2022. The Determinants of Consanguineous Marriages among the Arab Population: A Systematic Review. Iran J Public Health, 51, 253–265.

46. Elfatih, A., Saad, C., Biobank, 4, S. P. F. E. Q. F. A. E. A. N., Sequencing, 5, G. G. T. S. L. W. L. S., 6, A. B. C. S. N. A. H. V. F. R. T. R., Management, D., 7, C. I. G. S. T. A. K. M. H. H. M. Z. T. A. E. K. A. P. T. P. S. A.-A. R. & YOUNES 15 PUTHEN JITHESH V. 8 SUHRE KARSTEN 16 TATARI ZOHREH 17, C. L. P. I. A. O. M. A.-K. S. A. M. B. R. C. L. E. X. F. K. M. H. M. 2024. Analysis of 14,392 whole genomes reveals 3.5% of Qataris carry medically actionable variants. European Journal of Human Genetics, 32, 1465–1473.

47. Farnaes, L., Hildreth, A., Sweeney, N. M., Clark, M. M., Chowdhury, S., Nahas, S., Cakici, J. A., Benson, W., Kaplan, R. H. & Kronick, R. 2018. Rapid whole-genome sequencing decreases infant morbidity and cost of hospitalization. NPJ genomic medicine, 3, 10.

48. Fernandez-Marmiesse, A., Gouveia, S. & Couce, M. L. 2018. NGS technologies as a turning point in rare disease research, diagnosis and treatment. Current medicinal chemistry, 25, 404–432.

49. Ferreira, C. R. 2019. The burden of rare diseases. American Journal of Medical Genetics Part A, 179, 885–892.

50. Frederiksen, S. D., Avramović, V., Maroilley, T., Lehman, A., Arbour, L. & Tarailo-Graovac, M. 2022. Rare disorders have many faces: in silico characterization of rare disorder spectrum. Orphanet Journal of Rare Diseases, 17, 1–18.

51. Frésard, L. & Montgomery, S. B. 2018. Diagnosing rare diseases after the exome. Molecular Case Studies, 4, a003392.

52. Gambello, M. J. & Li, H. 2018. Current strategies for the treatment of inborn errors of metabolism. Journal of Genetics and Genomics, 45, 61–70.

53. Gentilini, A., Neez, E. & Wong-Rieger, D. 2025. Rare disease policy in high-income countries: an overview of achievements, challenges, and solutions. Value in Health.

54. Georgiou, T., Petrou, P. P., Malekkou, A., Ioannou, I., Gavatha, M., Skordis, N., Nicolaidou, P., Savvidou, I., Athanasiou, E. & Ourani, S. 2024. Inherited metabolic disorders in Cyprus. Molecular Genetics and Metabolism Reports, 39, 101083.

55. Ghanim, M., Mosleh, R., Hamdan, A., Amer, J., Alqub, M., Jarrar, Y. & Dwikat, M. 2023. Assessment of perceptions and predictors towards consanguinity: a cross-sectional study from Palestine. Journal of Multidisciplinary Healthcare, 3443–3453.

56. Glaubitz, R., Heinrich, L., Tesch, F., Seifert, M., Reber, K. C., Marschall, U., Schmitt, J. & Müller, G. 2025. The cost of the diagnostic odyssey of patients with suspected rare diseases. Orphanet Journal of Rare Diseases, 20, 222.

57. Gunaydin, M. & Bozkurter Cil, A. T. 2018. Progressive familial intrahepatic cholestasis: diagnosis, management, and treatment. Hepatic Medicine: Evidence and Research, 10, 95–104.

58. Haendel, M., Vasilevsky, N., Unni, D., Bologa, C., Harris, N., Rehm, H., Hamosh, A., Baynam, G., Groza, T. & Mcmurry, J. 2020. How many rare diseases are there? Nature reviews drug discovery, 19, 77–78.

59. Harari, S. 2016. Why we should care about ultra-rare disease. European Respiratory Review, 25, 101.

60. Hartley, T., Lemire, G., Kernohan, K. D., Howley, H. E., Adams, D. R. & Boycott, K. M. 2020. New diagnostic approaches for undiagnosed rare genetic diseases. Annual review of genomics and human genetics, 21, 351–372.

61. Iourov, I. Y., Vorsanova, S. G. & Yurov, Y. B. 2019. Pathway-based classification of genetic diseases. Molecular cytogenetics, 12, 1–5.

62. Janku, P., Robinow, M., Kelly, T., Bralley, R., Baynes, A., Edgerton, M. T. & Opitz, J. M. 1980. The van der Woude syndrome in a large kindred: variability, penetrance, genetic risks. American journal of medical genetics, 5, 117–123.

63. Jensson, B. O., Arnadottir, G. A., Katrinardottir, H., Fridriksdottir, R., Helgason, H., Oddsson, A., Sveinbjornsson, G., Eggertsson, H. P., Halldorsson, G. H. & Atlason, B. A. 2023. Actionable genotypes and their association with life span in Iceland. New England Journal of Medicine, 389, 1741–1752.

64. Kar, A., Sundaravadivel, P. & Dalal, A. 2024. Rare genetic diseases in India: Steps toward a nationwide mission program. Journal of Biosciences, 49, 34.

65. Katsanis, S. H. & Katsanis, N. 2013. Molecular genetic testing and the future of clinical genomics. Nature Reviews Genetics, 14, 415–426.

66. Kerdoncuff, E., Skov, L., Patterson, N., Banerjee, J., Khobragade, P., Chakrabarti, S. S., Chakrawarty, A., Chatterjee, P., Dhar, M., Gupta, M., John, J. P., Koul, P. A., Lehl, S. S., Mohanty, R. R., Padmaja, M., Perianayagam, A., Rajguru, C., Sankhe, L., Talukdar, A., Varghese, M., Yadati, S. R., Zhao, W., Leung, Y. Y., Schellenberg, G. D., Wang, Y. Z., Smith, J. A., Dey, S., Ganna, A., Dey, A. B., Kardia, S. L. R., Lee, J. & Moorjani, P. 2025. 50,000 years of evolutionary history of India: Impact on health and disease variation. Cell, 188, 3389–3404.e6.

67. Khalid, S. N., Midhet, F., Uzma, Q., Thom, E. M., Baqai, S., Khan, M. T. & Memon, A. 2025. Factors associated with induced abortions in Pakistan: a comprehensive analysis of Pakistan maternal mortality survey 2019. Frontiers in Reproductive Health, Volume 7 - 2025.

68. Khan, H. 2023. Rare bleeding disorders in Pakistan–a call for exploration! Pakistan Journal of Medical Sciences, 39.

69. Kingma, S. D. K. & Jonckheere, A. I. 2021. MPS I: Early diagnosis, bone disease and treatment, where are we now? Journal of Inherited Metabolic Disease, 44, 1289–1310.

70. Klimova, B., Storek, M., Valis, M. & Kuca, K. 2017. Global view on rare diseases: a mini review. Current medicinal chemistry, 24, 3153–3158.

71. Landoure, G., Samassekou, O., Traore, M., Meilleur, K. G., Guinto, C. O., Burnett, B. G., Sumner, C. J. & Fischbeck, K. H. 2016. Genetics and genomic medicine in Mali: challenges and future perspectives. Molecular genetics & genomic medicine, 4, 126.

72. Landrum, M. J., Chitipiralla, S., Brown, G. R., Chen, C., Gu, B., Hart, J., Hoffman, D., Jang, W., Kaur, K. & Liu, C. 2020. ClinVar: improvements to accessing data. Nucleic acids research, 48, D835–D844.

73. Lavelle, T. A., Feng, X., Keisler, M., Cohen, J. T., Neumann, P. J., Prichard, D., Schroeder, B. E., Salyakina, D., Espinal, P. S. & Weidner, S. B. 2022. Cost-effectiveness of exome and genome sequencing for children with rare and undiagnosed conditions. Genetics in Medicine, 24, 1349–1361.

74. Li, D., Tian, L. & Hakonarson, H. 2018. Increasing diagnostic yield by RNA-Sequencing in rare disease—bypass hurdles of interpreting intronic or splice-altering variants. Annals of Translational Medicine, 6.

75. Liascovich, R., Rittler, M. & Castilla, E. E. 2000. Consanguinity in South America: demographic aspects. Human Heredity, 51, 27–34.

76. Loomes, K. M., Squires, R. H., Kelly, D., Rajwal, S., Soufi, N., Lachaux, A., Jankowska, I., Mack, C., Setchell, K. D. & Karthikeyan, P. 2022. Maralixibat for the treatment of PFIC: Long-term, IBAT inhibition in an open-label, Phase 2 study. *Hepatology communications*, 6, 2379-2390.

77. Lorentsen, J. L. L., Brunborg, C., Aasebø, A. T., Valeur, J., Haavardsholm, E. A., Molberg, Ø., Palm, Ø. & Lerang, K. 2025. Incidence and prevalence of Behçet’s disease in Oslo, Norway: a two-decade population-based analysis. Rheumatology, keaf224.

78. Lu, Y., Gao, Q., Ren, X., Li, J., Yang, D., Zhang, Z. & Han, J. 2022. Incidence and prevalence of 121 rare diseases in China: current status and challenges: 2022 revision. Intractable & Rare Diseases Research, 11, 96–104.

79. Lumaka, A., Race, V., Peeters, H., Corveleyn, A., Coban-Akdemir, Z., Jhangiani, S. N., Song, X., Mubungu, G., Posey, J. & Lupski, J. R. 2018. A comprehensive clinical and genetic study in 127 patients with ID in Kinshasa, DR Congo. American Journal of Medical Genetics Part A, 176, 1897–1909.

80. Maher, T. M., Bendstrup, E., Dron, L., Langley, J., Smith, G., Khalid, J. M., Patel, H. & Kreuter, M. 2021. Global incidence and prevalence of idiopathic pulmonary fibrosis. Respiratory research, 22, 197.

81. Malik, S. & AMIN-UD-Din, M. 2013. Genetic heterogeneity and gene diversity at ABO and Rh loci in the human population of southern Punjab, Pakistan. Pakistan Journal of Zoology, 45.

82. Malinowski, J., Miller, D. T., Demmer, L., Gannon, J., Pereira, E. M., Schroeder, M. C., Scheuner, M. T., Tsai, A. C.-H., Hickey, S. E. & Shen, J. 2020. Systematic evidence-based review: outcomes from exome and genome sequencing for pediatric patients with congenital anomalies or intellectual disability. Genetics in Medicine, 22, 986–1004.

83. Manickam, K., Mcclain, M. R., Demmer, L. A., Biswas, S., Kearney, H. M., Malinowski, J., Massingham, L. J., Miller, D., Yu, T. W. & Hisama, F. M. 2021. Exome and genome sequencing for pediatric patients with congenital anomalies or intellectual disability: an evidence-based clinical guideline of the American College of Medical Genetics and Genomics (ACMG). Genetics in Medicine, 23, 2029–2037.

84. Mansouritorghabeh, H. 2021. Consanguineous marriage and rare bleeding disorders. Expert Review of Hematology, 14, 467–472.

85. Mariz, S., Reese, J. H., Westermark, K., Greene, L., Goto, T., Hoshino, T., Llinares-Garcia, J. & Sepodes, B. 2016. Worldwide collaboration for orphan drug designation. Nature reviews Drug discovery, 15, 440–441.

86. Marta, A., Marques, J. P., Santos, C., Coutinho-Santos, L., VAZ-Pereira, S., Costa, J., Arede, P., Félix, R., Geada, S. & Gouveia, N. 2024. The socioeconomic epidemiology of inherited retinal diseases in Portugal. Orphanet Journal of Rare Diseases, 19, 151.

87. Masri, A. T., Oweis, L., AL Qudah, A. & EL-Shanti, H. 2022. Congenital muscle dystrophies: Role of singleton whole exome sequencing in countries with limited resources. Clinical neurology and neurosurgery, 217, 107271.

88. Matencio, A., Navarro-Orcajada, S., González-Ramón, A., García-Carmona, F. & López-Nicolás, J. M. 2020. Recent advances in the treatment of Niemann pick disease type C: A mini-review. International Journal of Pharmaceutics, 584, 119440.

89. Mazzucato, M., VISONÀ DALLA Pozza, L., Minichiello, C., Toto, E., Vianello, A. & Facchin, P. 2023. Estimating mortality in rare diseases using a population-based registry, 2002 through 2019. Orphanet Journal of Rare Diseases, 18, 362.

90. Mccullough, J. M. & O’rourke, D. H. 1986. Geographic distribution of consanguinity in Europe. Annals of Human Biology, 13, 359–367.

91. Mckiernan, P., Bernabeu, J. Q., Girard, M., Indolfi, G., Lurz, E. & Trivedi, P. 2024. Opinion paper on the diagnosis and treatment of progressive familial intrahepatic cholestasis. JHEP Reports, 6, 100949.

92. Merker, J. D., Wenger, A. M., Sneddon, T., Grove, M., Zappala, Z., Fresard, L., Waggott, D., Utiramerur, S., Hou, Y. & Smith, K. S. 2018. Long-read genome sequencing identifies causal structural variation in a Mendelian disease. Genetics in Medicine, 20, 159–163.

93. Mizuguchi, T., Suzuki, T., Abe, C., Umemura, A., Tokunaga, K., Kawai, Y., Nakamura, M., Nagasaki, M., Kinoshita, K. & Okamura, Y. 2019. A 12-kb structural variation in progressive myoclonic epilepsy was newly identified by long-read whole-genome sequencing. Journal of human genetics, 64, 359–368.

94. Modell, B., Darlison, M., Moorthie, S., Blencowe, H., Petrou, M. & Lawn, J. 2016. Epidemiological methods in community genetics and the Modell Global Database of Congenital Disorders (MGDb).

95. Mofied, E. A., Abo-Elkheir, O. I., Gaber, K. R. & Abd EL Fattah, T. A. 2024. The effect of consanguineous marriage on reproductive wastage and Perinatal outcomes. Journal of Recent Advances in Medicine, 5, 135–141.

96. Mogess, W. N. & Mihretie, T. B. 2024. Prevalence and associated factors of congenital anomalies in Ethiopia: A systematic review and meta-analysis. Plos one, 19, e0302393.

97. Mohyuddin, A., Ayub, Q., Khaliq, S., Mansoor, A., Mazhar, K., Rehman, S. & Mehdi, S. Q. 2002. HLA polymorphism in six ethnic groups from Pakistan. Tissue Antigens, 59, 492–501.

98. Moliner, A. M. & Waligora, J. 2017. The European Union policy in the field of rare diseases. Rare diseases epidemiology: Update and overview. Springer.

99. Naqvi, S. F., Ameena, U., Qazi, W. U., Ahmad, S., Iqbal, A. & Malik, S. 2024. Burden of congenital and hereditary anomalies and their epidemiological attributes in the pediatric and adult population of Peshawar valley, Pakistan. Pak J Med Sci, 40, 2181–2189.

100. Naumova, E., Lesichkova, S., Milenova, V., Yankova, P., Murdjeva, M. & Mihailova, S. 2022. Primary immunodeficiencies in Bulgaria-achievements and challenges of the PID National Expert Center. Frontiers in immunology, 13, 922752.

101. Nawaz, A., Siddiqui, A., Mughal, M., Naz, S., Wajid, M. & Malik, S. 2025. Congenital anomalies in Okara District of Pakistan: Epidemiology, spectrum and ethno-demographic inequalities. Pak J Med Sci, 41, 643–651.

102. Nguengang Wakap, S., Lambert, D. M., Olry, A., Rodwell, C., Gueydan, C., Lanneau, V., Murphy, D., LE Cam, Y. & Rath, A. 2020. Estimating cumulative point prevalence of rare diseases: analysis of the Orphanet database. European Journal of Human Genetics, 28, 165–173.

103. Nguyen, T. T., To, H. T. T., Le, A. N. T., Pham, A. Q., Nguyen, N. D., Ha, H. H., Vu, H. T., Hoang, T. T. & Tran, M. C. 2025. Prevalence of common autosomal recessive and X-linked conditions in pregnant women in Vietnam: a cross-sectional study. Scientific Reports, 15, 19054.

104. Obeidat, L., Abu-Halaweh, M., Alzyoud, R., Albsoul, E. & Zaravinos, A. 2024. Genetic causes of primary immunodeficiency in the Jordanian population. Biomedical Reports, 21, 160.

105. Owen, M. J., Niemi, A.-K., Dimmock, D. P., Speziale, M., Nespeca, M., Chau, K. K., Van der Kraan, L., Wright, M. S., Hansen, C. & Veeraraghavan, N. 2021. Rapid sequencing-based diagnosis of thiamine metabolism dysfunction syndrome. New England Journal of Medicine, 384, 2159–2161.

106. Passos-Bueno, M. R., Bertola, D., Horovitz, D. D. G., De Faria Ferraz, V. E. & Brito, L. A. 2014. Genetics and genomics in Brazil: a promising future. Molecular genetics & genomic medicine, 2, 280–291.

107. Pathak, A. K., Kadian, A., Kushniarevich, A., Montinaro, F., Mondal, M., Ongaro, L., Singh, M., Kumar, P., Rai, N., Parik, J., Metspalu, E., Rootsi, S., Pagani, L., Kivisild, T., Metspalu, M., Chaubey, G. & Villems, R. 2018. The Genetic Ancestry of Modern Indus Valley Populations from Northwest India. The American Journal of Human Genetics, 103, 918–929.

108. Perolla, A., Çalliku, E., Cili, A., Caja, T., Pulluqi, P. & Ivanaj, A. 2024. Uncovering the Challenges of Rare Diseases: Insights From a Retrospective Cross-Sectional Study in Albania (2005-2022). Cureus, 16.

109. Plaiasu, V., Nanu, M. & Matei, D. 2010. Rare Disease Day–at a glance. Maedica, 5, 65.

110. Popescu, G., Rusu, C., Maștaleru, A., Oancea, A., Cumpăt, C. M., Luca, M. C., Grosu, C. & Leon, M. M. 2025. Social and Demographic Determinants of Consanguineous Marriage: Insights from a Literature Review. Genealogy, 9, 69.

111. Richards, S., Aziz, N., Bale, S., Bick, D., Das, S., GastieR-Foster, J., Grody, W. W., Hegde, M., Lyon, E. & Spector, E. 2015. Standards and guidelines for the interpretation of sequence variants: a joint consensus recommendation of the American College of Medical Genetics and Genomics and the Association for Molecular Pathology. Genetics in medicine, 17, 405–423.

112. Richter, T., Nestler-Parr, S., Babela, R., Khan, Z. M., Tesoro, T., Molsen, E. & Hughes, D. A. 2015. Rare disease terminology and definitions—a systematic global review: report of the ISPOR rare disease special interest group. Value in health, 18, 906–914.

113. Rode, J. 2021. Rare diseases: understanding this public health priority. 2005.

114. Romdhane, L., Mezzi, N., Hamdi, Y., EL-Kamah, G., Barakat, A. & Abdelhak, S. 2019. Consanguinity and inbreeding in health and disease in North African populations. Annual review of genomics and human genetics, 20, 155–179.

115. SAJID Hussain, M., MARRIAM Bakhtiar, S., Farooq, M., Anjum, I., Janzen, E., REZA Toliat, M., Eiberg, H., Kjaer, K., Tommerup, N. & Noegel, A. 2013. Genetic heterogeneity in Pakistani microcephaly families. Clinical genetics, 83, 446–451.

116. Sanford, E. F., Clark, M. M., Farnaes, L., Williams, M. R., Perry, J. C., Ingulli, E. G., Sweeney, N. M., Doshi, A., Gold, J. J. & Briggs, B. 2019. Rapid whole genome sequencing has clinical utility in children in the PICU. Pediatric Critical Care Medicine, 20, 1007–1020.

117. SANTOS-Lozano, A., VILLAMANDOS García, D., SANCHIS-Gomar, F., FIUZA-Luces, C., PAREJA-Galeano, H., Garatachea, N., NOGALES Gadea, G. & Lucia, A. 2015. Niemann-Pick disease treatment: a systematic review of clinical trials. Ann Transl Med, 3, 360.

118. Sathar, Z., Singh, S., Shah, I. H., Niazi, M. R., Parveen, T., Mulhern, O. & Mir, A. M. 2025. Abortion and unintended pregnancy in Pakistan: new evidence for 2023 and trends over the past decade. BMJ Global Health, 10, e017239.

119. Sathar, Z. A., Singh, S. & Fikree, F. F. 2007. Estimating the Incidence of Abortion in Pakistan. Studies in Family Planning, 38, 11–22.

120. Sciascia, S., Roccatello, D., Salvatore, M., Carta, C., Cellai, L. L., Ferrari, G., Lumaka, A., Groft, S., Alanay, Y. & Azam, M. 2023. Unmet needs in countries participating in the undiagnosed diseases network international: an international survey considering national health care and economic indicators. Frontiers in Public Health, 11, 1248260.

121. Sequeira, A. R., Mentzakis, E., Archangelidi, O. & Paolucci, F. 2021. The economic and health impact of rare diseases: A meta-analysis. Health Policy and Technology, 10, 32–44.

122. Shafie, A. A., Supian, A., Ahmad Hassali, M. A., Ngu, L.-H., Thong, M.-K., Ayob, H. & Chaiyakunapruk, N. 2020. Rare disease in Malaysia: Challenges and solutions. PLoS One, 15, e0230850.

123. Sharma, A., Jacob, A., Tandon, M. & Kumar, D. 2010. Orphan drug: development trends and strategies. Journal of Pharmacy and Bioallied Sciences, 2, 290–299.

124. Shenk, M. K., Naz, S. & Chaudhry, T. 2024. Intensive kinship, development, and demography: Why Pakistan has the highest rates of cousin marriage in the world. Population and Development Review, 50, 1045–1090.

125. Sherry, S. T., Ward, M. & Sirotkin, K. 1999. dbSNP-database for single nucleotide polymorphisms and other classes of minor genetic variation. Genome Research, 9, 677–9.

126. Sihombing, N. R. B., Winarni, T. I., Utari, A., VAN Bokhoven, H., Hagerman, R. J. & Faradz, S. M. 2021. Surveillance and prevalence of fragile X syndrome in Indonesia. Intractable & Rare Diseases Research, 10, 11–16.

127. Sirisena, N. D. & Dissanayake, V. H. 2019. Genetics and genomic medicine in Sri Lanka. Molecular Genetics & Genomic Medicine, 7, e744.

128. Small, N., Bittles, A. H., Petherick, E. S. & Wright, J. 2017. Endogamy, consanguinity and the health implications of changing marital choices in the UK Pakistani community. Journal of biosocial science, 49, 435–446.

129. Smith, C. E., Bergman, P. & Hagey, D. W. 2022. Estimating the number of diseases–the concept of rare, ultra-rare, and hyper-rare. Iscience, 25.

130. Song, P., Gao, J., Inagaki, Y., Kokudo, N. & Tang, W. 2012. Rare diseases, orphan drugs, and their regulation in Asia: Current status and future perspectives. Intractable & rare diseases research, 1, 3–9.

131. Stapleton, M., Hoshina, H., Sawamoto, K., Kubaski, F., Mason, R. W., Mackenzie, W. G., Theroux, M., Kobayashi, H., Yamaguchi, S., Suzuki, Y., Fukao, T., Tadao, O., Ida, H. & Tomatsu, S. 2019. Critical review of current MPS guidelines and management. Molecular Genetics and Metabolism, 126, 238–245.

132. Tarailo-Graovac, M., Shyr, C., Ross, C. J., Horvath, G. A., Salvarinova, R., Ye, X. C., Zhang, L.-H., Bhavsar, A. P., Lee, J. J. & Drögemöller, B. I. 2016. Exome sequencing and the management of neurometabolic disorders. New England Journal of Medicine, 374, 2246–2255.

133. Theadom, A., Rodrigues, M., Poke, G., O’grady, G., Love, D., Hammond-Tooke, G., Parmar, P., Baker, R., Feigin, V. & Jones, K. 2019. A nationwide, population-based prevalence study of genetic muscle disorders. Neuroepidemiology, 52, 128–135.

134. Thevenon, J., Duffourd, Y., Masurel-Paulet, A., Lefebvre, M., Feillet, F., El Chehadeh-Djebbar, S., ST-Onge, J., Steinmetz, A., Huet, F. & Chouchane, M. 2016. Diagnostic odyssey in severe neurodevelopmental disorders: toward clinical whole-exome sequencing as a first-line diagnostic test. Clinical Genetics, 89, 700–707.

135. Tiivoja, E., Reinson, K., Muru, K., Rähn, K., Muhu, K., Mauring, L., Kahre, T., Pajusalu, S. & Õunap, K. 2022. The prevalence of inherited metabolic disorders in Estonian population over 30 years: A significant increase during study period. JIMD reports, 63, 604–613.

136. Tournebize, R., Chu, G. & Moorjani, P. 2022. Reconstructing the history of founder events using genome-wide patterns of allele sharing across individuals. PLOS Genetics, 18, e1010243.

137. Venugopal, N., Naik, G., Jayanna, K., Mohapatra, A., Sasinowski, F. J., Kartha, R. V. & Rajasimha, H. K. 2024. Review of methods for estimating the prevalence of rare diseases. Rare Disease and Orphan Drugs Journal, 3, N/A-N/A.

138. Wainstock, D. & Katz, A. 2023. Advancing rare disease policy in Latin America: a call to action. The Lancet Regional Health–Americas, 18.

139. Wang, Q., Lu, Q. & Zhao, H. 2015. A review of study designs and statistical methods for genomic epidemiology studies using next generation sequencing. Frontiers in genetics, 6, 149.

140. Warsy, A. S., AL-Jaser, M. H., Albdass, A., AL-Daihan, S. & Alanazi, M. 2014. Is consanguinity prevalence decreasing in Saudis?: A study in two generations. African health sciences, 14, 314–321.

141. Wasim, M., Khan, H. N., Ayesha, H. & Awan, F. R. 2023. Need and Challenges in Establishing Newborn Screening Programs for Inherited Metabolic Disorders in Developing Countries. Advanced Biology, 2200318.

142. Wehrli, S., Dwyer, A. A., Baumgartner, M. R., Lehmann, C. & Landolt, M. A. 2024. Lower Healthcare Access and Its Association With Individual Factors and Health-Related Quality of Life in Adults With Rare Diseases in Switzerland. International Journal of Public Health, 69, 1607548.

143. Wenger, A. M., Guturu, H., Bernstein, J. A. & Bejerano, G. 2017. Systematic reanalysis of clinical exome data yields additional diagnoses: implications for providers. Genetics in Medicine, 19, 209–214.

144. Willig, L. K., Petrikin, J. E., Smith, L. D., Saunders, C. J., Thiffault, I., Miller, N. A., Soden, S. E., Cakici, J. A., Herd, S. M. & Twist, G. 2015. Whole-genome sequencing for identification of Mendelian disorders in critically ill infants: a retrospective analysis of diagnostic and clinical findings. The Lancet Respiratory Medicine, 3, 377–387.

145. Witzel, M. 2019. Early ‘Aryans’ and their neighbors outside and inside India. Journal of biosciences, 44, 1–10.

146. Woerner, A. C., Gallagher, R. C., Vockley, J. & Adhikari, A. N. 2021. The use of whole genome and exome sequencing for newborn screening: challenges and opportunities for population health. Frontiers in Pediatrics, 9, 652.

147. Yu, E., Ambati, A., Andersen, M. S., Krohn, L., Estiar, M. A., Saini, P., Senkevich, K., Sosero, Y. L., Sreelatha, A. A. K. & Ruskey, J. A. 2021. Fine mapping of the HLA locus in Parkinson’s disease in Europeans. npj Parkinson’s Disease, 7, 84.

148. Zaman, Q., Alharthi, M. T. H., Alahmari, S. A. S., Khan, T., Abbas, M., Latif, M. & Jelani, M. 2025. Whole exome sequencing: Unlocking the molecular diagnostic odyssey in Pakhtun ethnic group of Pakistani population. Gene, 962, 149586.

149. Zambrano-Mila, M. S., Agathos, S. N. & Reichardt, J. K. 2019. Human genetics and genomics research in Ecuador: historical survey, current state, and future directions. Human genomics, 13, 64.

150. Zar, M. S., Akhtar, M. S., Haris, A. R. & Aslamkhan, M. 2020. Colour Vision Deficiency and Consanguinity in Pakistani Pukhtoon Population. Advancements in Life Sciences, 7, 237–239.

